# Spatially-Resolved Transcriptomics Define Clinically Relevant Subsets of Macrophages in Diffuse Large B-cell Lymphoma

**DOI:** 10.1101/2023.03.05.23286445

**Authors:** Min Liu, Giorgio Bertolazzi, Kevin Mulder, Shruti Sridhar, Rui Xue Lee, Patrick Jaynes, Michal Marek Hoppe, Shuangyi Fan, Yanfen Peng, Jocelyn Thng, Reiya Chua, Sanjay De Mel, Limei Poon, Esther Chan, Joanne Lee, Susan Swee-Shan Hue, Siok-Bian Ng, K George Chandy, Florent Ginhoux, Yen Lin Chee, Claudio Tripodo, Anand D. Jeyasekharan

## Abstract

**Background:** Macrophages are abundant immune cells in the microenvironment of diffuse large B-cell lymphoma (DLBCL). Conventional immunohistochemistry-based studies with varying prognostic significance precludes a comprehensive analysis of macrophage subtypes in DLBCL. We hypothesized that whole-transcriptomic analysis (WTA) of macrophage in-situ would identify new macrophage subsets of biological and clinical significances.

**Methods:** Digital spatial profiling with WTA of CD68+ cells was performed in 47 DLBCL and 17 reactive lymphoid tissues (RLTs), to define macrophage signatures (termed “MacroSigs”) of distinct lymphoid spatial niches and clinical scenarios. Eight independent DLBCL datasets (4,594 patients) with transcriptomic and survival information were used for validation of MacroSigs.

**Results:** Digital spatial profiling revealed previously unrecognized transcriptomic differences between macrophages populating distinct spatial compartments in RLTs (light zone (LZ)/ dark zone (DZ), germinal center (GC)/ interfollicular (IF) regions), and in between disease states (RLTs and DLBCL with or without relapsed disease). This transcriptomic diversity of macrophages was categorized into eight MacroSigs. Spatial-MacroSigs associate with specific cell-of-origin (COO) subtypes of DLBCL, of particular interest being the IF-MacroSig enriched in the unclassified COO (*P*<0.005, 6/8 datasets). MacroSigs of relapsed-DLBCL and DZ were prognostic for shorter overall survival (*P* <0.05 in 5/8 datasets; *P* <0.05 in 8/8 datasets, respectively). Projection onto a macrophage single-cell RNA-sequencing atlas reveals the Non-relapse-DLBCL MacroSig to depict HES1/FOLR2-like macrophages, while relapse-DLBCL-MacroSig represents IL1B-like monocytes, with unique therapeutic vulnerabilities for each.

**Conclusions:** This study first provides spatially-resolved macrophage WTA in reactive and malignant lymphoid tissues. Gene expression signatures of macrophages in the DZ and relapsed-DLBCL samples are consistently prognostic in multiple datasets and offer insights into novel therapeutic strategies for DLBCL.

## Introduction

Diffuse large B-cell lymphoma (DLBCL) is the most common subtype of non-Hodgkin lymphoma in adults (1, 2). Combination therapy of rituximab with cyclophosphamide, doxorubicin, vincristine, and prednisolone (R-CHOP) is potentially curative, but 30%-40% of cases relapse after initial therapy (3). Elucidating mechanisms underlying relapse following R-CHOP is critical for the development of novel therapeutic strategies to improve DLBCL outcomes. An important and emerging area is the role of the tumour microenvironment (TME) in mediating disease progression and clearance of tumor cells after chemotherapy (4–8). Understanding factors in the TME that mediate DLBCL relapse following R-CHOP is also important in terms of incorporating novel immunotherapeutics such as anti-CD47 antibodies, bispecific CD20-CD3 T-cell engagers and CD19 CAR-T cells into front-line treatment regimens for DLBCL.

Tumor-associated macrophages (TAMs) are among the most abundant immune cells in DLBCL TME. TAMs are recognized as potential therapeutic targets in oncology due to driving tumor progression, metastasis, and recurrence (9). In DLBCL, TAM infiltration is associated with poor prognosis after R-CHOP therapy (7, 10, 11). However, there are discrepancies observed between studies, with a lack of sufficient reproducibility to identify consistent clinical prognostic markers (12). These discrepancies may partly result from the simplified functional classification of macrophages into “M1/ M2” phenotypes (13). Conventionally, M1 macrophages refer to a pro-inflammatory phenotype with pathogen-killing abilities while M2 refers to an anti-inflammatory phenotype promoting proliferation, tissue repair and tumorigenesis (14). However, macrophages in several physiological or pathological settings, including embryonic macrophages, resolution-phase macrophages and even certain TAMs do not clearly fall into either the M1 or M2 phenotype (15). Furthermore, macrophages show high phenotypic plasticity (15), suggesting that the M1/ M2 dichotomy does not fully encompass the functional diversity of macrophages (15–17). Indeed, with single-cell transcriptomic approaches, the functional complexity of macrophages is now widely appreciated (18, 19). The nature of TAMs defined by comprehensive transcriptomic approaches in DLBCL and their relationship with treatment outcomes remain poorly understood (20).

In this study, we aimed to comprehensively characterize TAMs in DLBCL and reactive lymphoid tissues (RLTs) using digital spatial profiling (DSP), an advanced technique for spatially resolved transcriptomics. The DSP whole transcriptome atlas (WTA) provides an unbiased map of 18000+ RNA targets throughout specific cell types of interest (chosen based on a fluorescent protein marker-here CD68 for macrophages) in formalin-fixed paraffin-embedded tissue sections. This allows characterization of macrophage biology within distinct spatial areas of RLTs, and in distinct DLBCL clinical subsets (patients with and without relapse after R-CHOP).

## Results

### DSP identifies transcriptomic patterns from distinct cell types within lymphoid tissue

We profiled the whole transcriptome of macrophages, T cells, and B cells based on the masks generated by immunofluorescent staining of the morphology markers CD68, CD3 and CD20, from spatial compartments in RLTs (*N* = 17) and DLBCL patients (*N* = 47) using the GeoMx® DSP WTA assay (Figure 1A) for a total of 252 Areas of Interest (AOIs) (Supplementary Figure S1A, representative images in Figure 1B-C and Supplementary Figure S1B, quality control and normalization in Supplementary Figure S1C).

**Figure 1.**
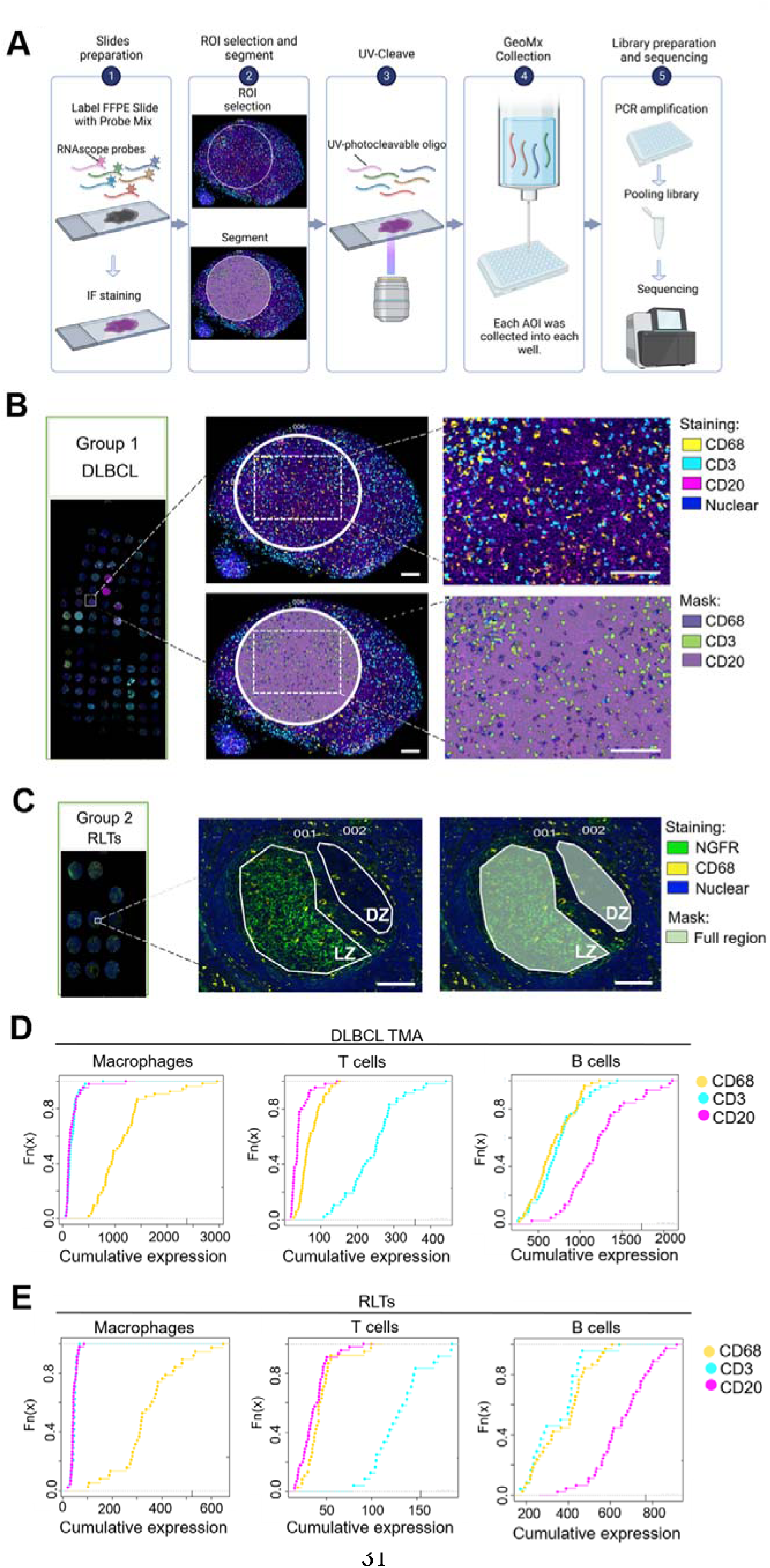
DSP identifies transcriptomic patterns from distinct cell types within lymphoid tissue. **(A)** Schematic of GeoMx® DSP WTA workflow. Immunofluorescence staining of DLBCL tissues **(B)** and RLTs **(C)**. In Group 1, CD68 stained macrophages (yellow), CD3 stained T cells (cyan), CD20 stained B cells (magenta), and SYTO 13 stained nuclei (blue). In Group 2, CD68 stained macrophages (yellow), NGFR illuminated LZ (green) and SYTO 13 stains nuclei (blue). After ROIs selection, each cell type was segmented by their corresponding masks. Representative images were shown. Scale bar: 100 μm. **(D-E)** Cumulative density functions showed that the signatures of macrophage, T cell, and B cell were highly enriched in CD68+ regions, CD3+ regions, and CD20+ regions, respectively in RLTs and DLBCL tissues. reactive lymphoid tissues, RLTs; diffuse large B-cell lymphoma, DLBCL; regions of interest, ROIs; areas of interest, AOIs; formalin-fixed paraffin-embedded, FFPE; light zone, LZ; dark zone, DZ; germinal center, GC; nerve growth factor receptor, NGFR.

Given the heterogeneity and high cellularity of the DLBCL microenvironment, we first wished to confirm that DSP-based cell selection generated reliable cell-type specific data. We therefore cross-validated DSP output with prior scRNA-seq data of RLTs and DLBCL specimens. We first defined gene signatures characteristic of macrophages, T cells and B cells using key genes known to be expressed in these cell types (Supplementary Table S2). We found that these gene-signatures closely overlapped with matching cell-types on scRNA-seq (Supplementary Figure S2). We then tested these signatures on our DSP based CD68+, CD20+ and CD3+ AOIs using cumulative density function. As expected, the macrophage signature was highly enriched in the CD68+ AOIs, and the signatures of T and B cells were highly enriched in CD3+ and CD20+ AOIs, respectively (Figure 1D-E, *P* < 0.05, Kolmogorov-Smirnov test,). This provides evidence that the morphology marker based AOI accurately captures the respective cell type of interest.

We further evaluated the robustness of our DSP platform by cross-comparing its output for differentially expressed genes (DEGs) between light zone (LZ) and dark zone (DZ) (Supplementary Figure S3A-B) with previously published transcriptomic differences between B-cells in the LZ and DZ microenvironments in the germinal center (GC) of RLTs (21–23). Almost all DEGs of CD20-highlighted cells in our DSP experiment overlapped with previously reported B-cell signatures (23) of LZ and DZ (Supplementary Figure S3C), providing evidence of adequate transcriptomic coverage for subsequent analyses and inferences.

### Unique gene expression patterns differentiate macrophages in the GC and in interfollicular (IF) areas of RLTs

Having quality checked the DSP data, we next aimed to investigate transcriptomic differences of CD68+ macrophages within different spatial regions of RLTs. An unsupervised clustering analysis highlighted sharp GC and IF separation (Figure 2A), suggesting highly distinct gene expression patterns. On further DEG analysis, we observed that 792 and 1036 genes were upregulated in the GC and IF macrophages respectively (Figure 2B-C, *P* < 0.05, Moderated *t*-test). Notably, macrophages in the IF showed upregulation of S100A family members (calcium binding proteins), such as S100A2, S100A8, and S100A9, previously reported to be high in macrophages that experience stimuli-induced M2-like differentiation (24). Enriched pathways in GC macrophages were mostly associated with MYC transcriptional regulation, E2F targets, and G2M checkpoint (*P* < 0.0001, Fisher’s exact test) (Figure 2D). In contrast, enriched pathways in IF were mostly associated with immune responses such as interferon response, Tumor necrosis factor (TNF)-α inflammatory response, and the Complement system (Figure 2D, *P* < 0.0005, Fisher’s exact test). The GC consists of two functionally distinct compartments: DZ and LZ (25). This compartmentalization is critical for dynamic differentiation of B cells within the GC (26). Therefore, we also compared the DEGs of macrophages in the LZ and DZ. Several genes and pathways were distinctly expressed between LZ and DZ macrophages, albeit not statistically significant at 0.05 for corrected p-values (Figure 2E-G, Moderated *t*-test). Based on these DEGs between GC versus IF, and LZ versus DZ, we derived MacroSigs corresponding to spatial compartments in reactive lymphoid tissue (RLT): defined henceforth as MacroSig1 (GC), MacroSig2 (IF), MacroSig3 (LZ) and MacroSig4 (DZ). We then evaluated if these spatially-derived macrophage DEGs could be mapped to known macrophage subclusters. For mapping, we utilized an integrative dataset named “MoMac-VERSE” (27) (Figure 2H), a large combined analysis of human monocytes and macrophages, with 17 annotated monocyte/ macrophage subclusters from 41 scRNA-seq datasets comprising healthy and cancer tissues. When we projected the top 50 genes or all genes of our MacroSigs (Supplementary Table S3) onto MoMac-VERSE, neither MacroSig1 (GC) nor MacroSig2 (IF) seemed to overlay with a specific subcluster of macrophages (Figure 2I-L). The distribution of MacroSig3 (LZ) in the MoMac-VERSE roughly project to CD16-monocytes (Figure 2M-N), while MacroSig4 (DZ) seemingly overlap with HES1/FOLR2 macrophages (Figure 2O-P). HES1/FOLR2 macrophages have been observed in many cancer-related studies and are considered to be tissue-resident macrophages (TRMs) that were associated with a poor clinical outcome (28, 29). Overall, these findings indicate that macrophages in different spatial locations of RLTs have different biological characteristics (with GC/IF macrophages representing a hitherto unknown macrophage subset).

**Figure 2.**
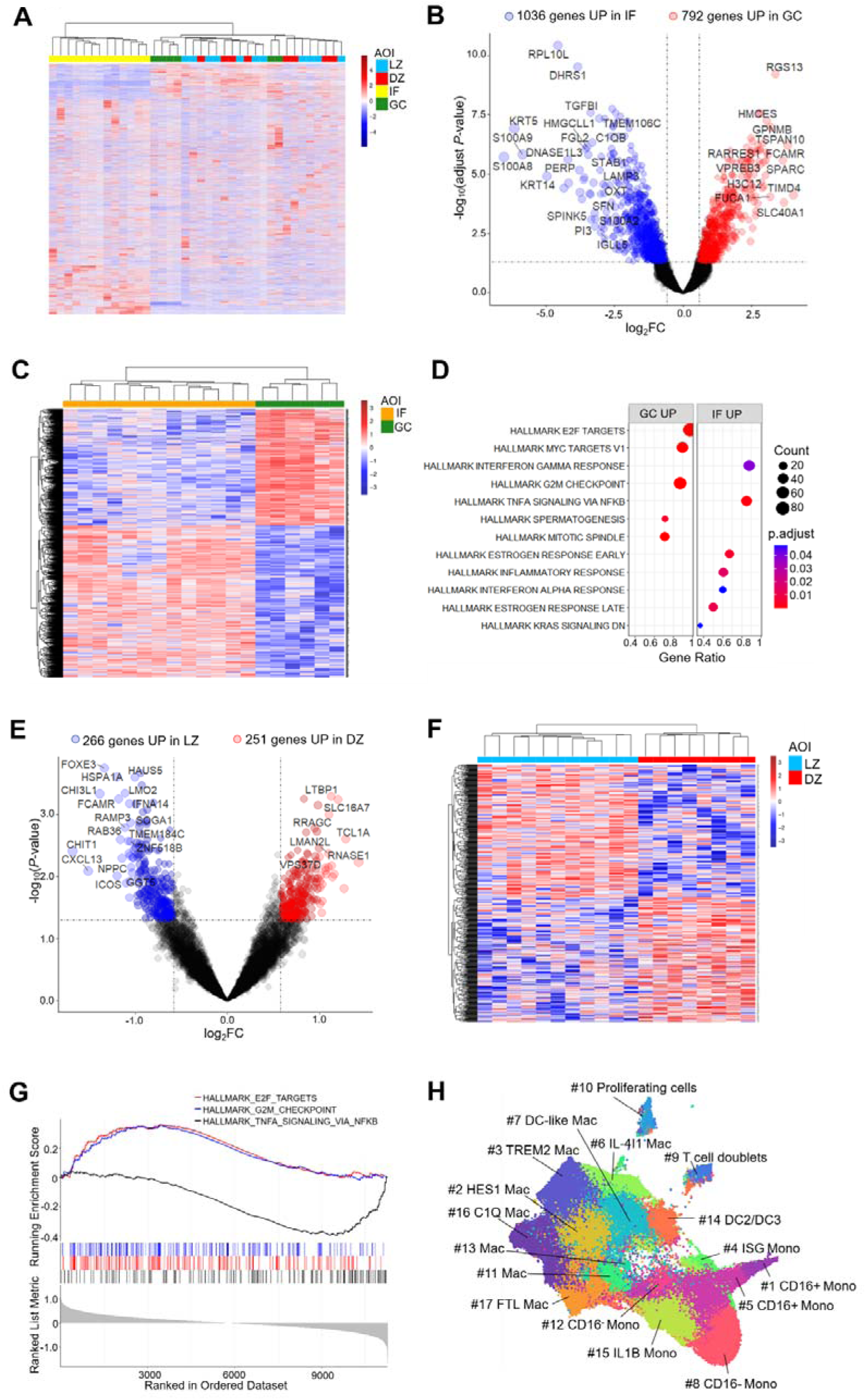
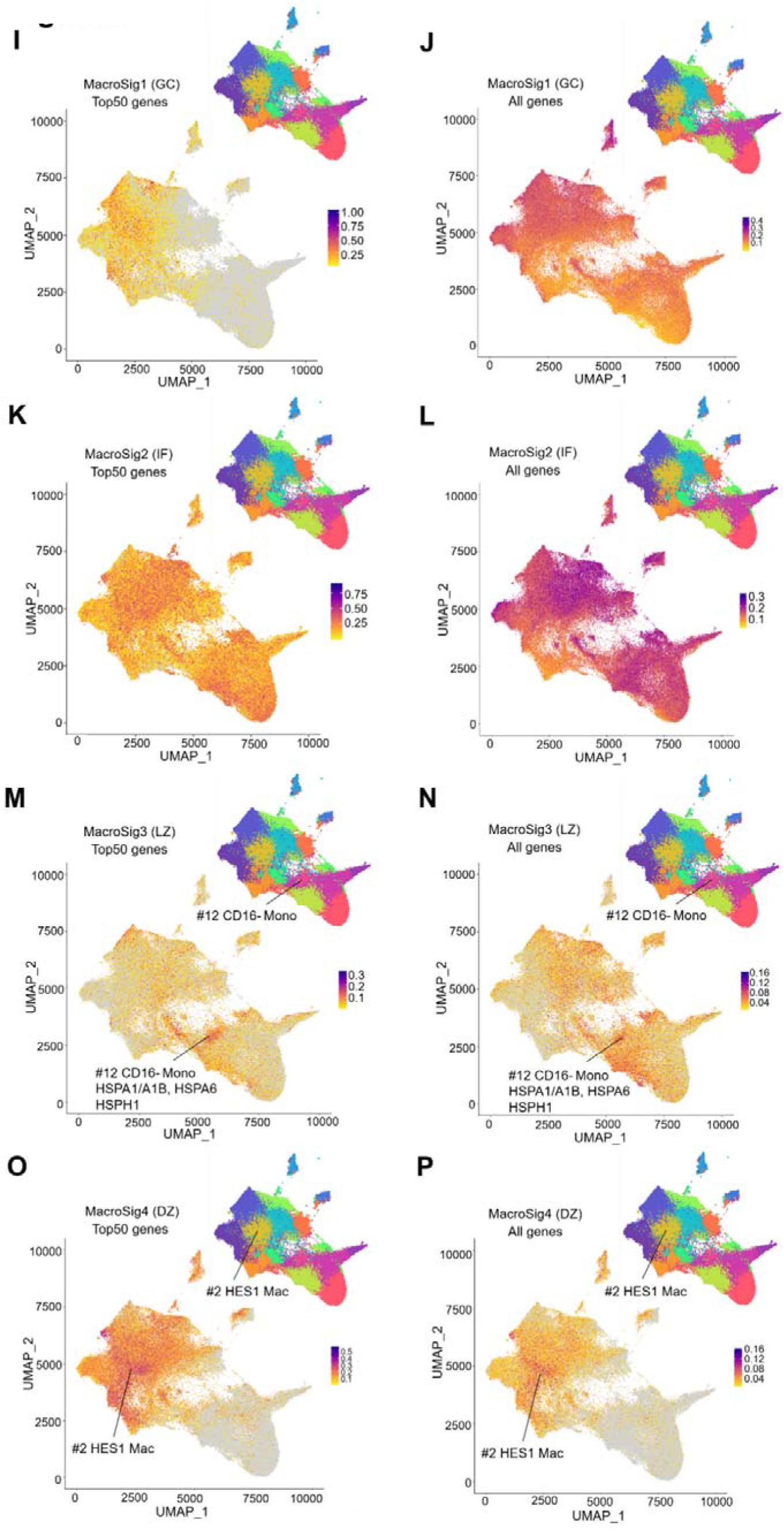
Unique gene expression patterns differentiate macrophages in the GC and interfollicular (IF) areas of RLTs. **(A)** Unsupervised clustering analysis was performed on transcripts from CD68+ masks within different spatial localizations of RLTs. **(B-C).** The volcano plot and heatmap showed the macrophage DEGs between GC and IF based on adjusted *P*-value (p<0.05, |log_2_-FC| ≥0.58). **(D)** GSEA was performed on the significant macrophage DEGs between GC and IF (p<0.05, |log_2_-FC|≥0.58). **(E-F).** The volcano plot and heatmap showed the macrophage DEGs between LZ and DZ based on *P*-value (p<0.05, |log_2_-FC|≥0.58). **(G)** GSEA was performed on all macrophage DEGs between LZ and DZ. The top three enriched signaling pathway were displayed. **(H)** MoMac-VERSE annotated 17 TAM subclusters using a compilation of 41 scRNA-seq datasets from healthy and cancer tissues. **(I-L)** MacroSig1 (GC) and MacroSig2 (IF) were projected into MoMac-VERSE, respectively. **(M-P)** MacroSig3 (LZ) and MacroSig4 (DZ) were projected into MoMac-VERSE, respectively. fold change, FC; gene set enrichment analysis, GSEA; macrophage signatures, MacroSigs; monocytes, Mono.

### Distinct transcriptomic profiles of macrophages in RLTs and DLBCL

DLBCL is thought to originate from different functional stages of GC B cells (30), and thus macrophages from RLT GC regions were used as the comparator for macrophages from DLBCL samples. An unsupervised clustering analysis using genes derived from CD68+ masks of RLTs GC regions and DLBCL samples revealed distinct subgroups corresponding to these two sets (Figure 3A), referred to henceforth as MacroSig5 (RLT) and MacroSig6 (DLBCL). To determine specific differences, we investigated the DEGs of these two macrophage subsets; 183 and 176 unique genes were highly expressed in RLTs GC and DLBCL macrophages, respectively (Figure 3B-C, *P* < 0.05, Moderated *t*-test). CD163, a marker of pro-tumorigenic macrophages, was high in DLBCL DEGs (Figure 3B, *P* = 0.01, Moderated *t*-test). The Complement pattern recognition components C1QA, C1QB, and C1QC were also significantly upregulated in DLBCL macrophages (Figure 3B, *P* < 0.0001, Moderated *t*-test), as were the entire Hallmark gene set of the Complement pathway (Figure 3D, *P* < 0.0001, Fisher’s exact test). This suggests a possible role for the Complement system in macrophage polarization in the DLBCL TME. Furthermore, the most enriched signaling pathway in DLBCL macrophages was TNF-α signaling via NF-κB (*P* < 0.0001, Fisher’s exact test). Finally, we projected MacroSig5 (RLT) and MacroSig6 (DLBCL) onto the MoMac-VERSE. While the MacroSig5 (RLT) did not specify a population (Figure 3E-F), the MacroSig6 (DLBCL) overlapped with the IL4I1 macrophage population (Figure 3G-H), which embodies an immunoregulatory function associated with tumors (27, 31, 32). In conjunction with the enrichment of the Complement and TNF-α signaling pathways, these results show that IL4I1-type DLBCL TAMs have a rewired transcriptional landscape distinct from that of macrophages populating RLTs GCs, and provides a starting point for work to evaluate their mechanistic role in DLBCL evolution.

**Figure 3.**
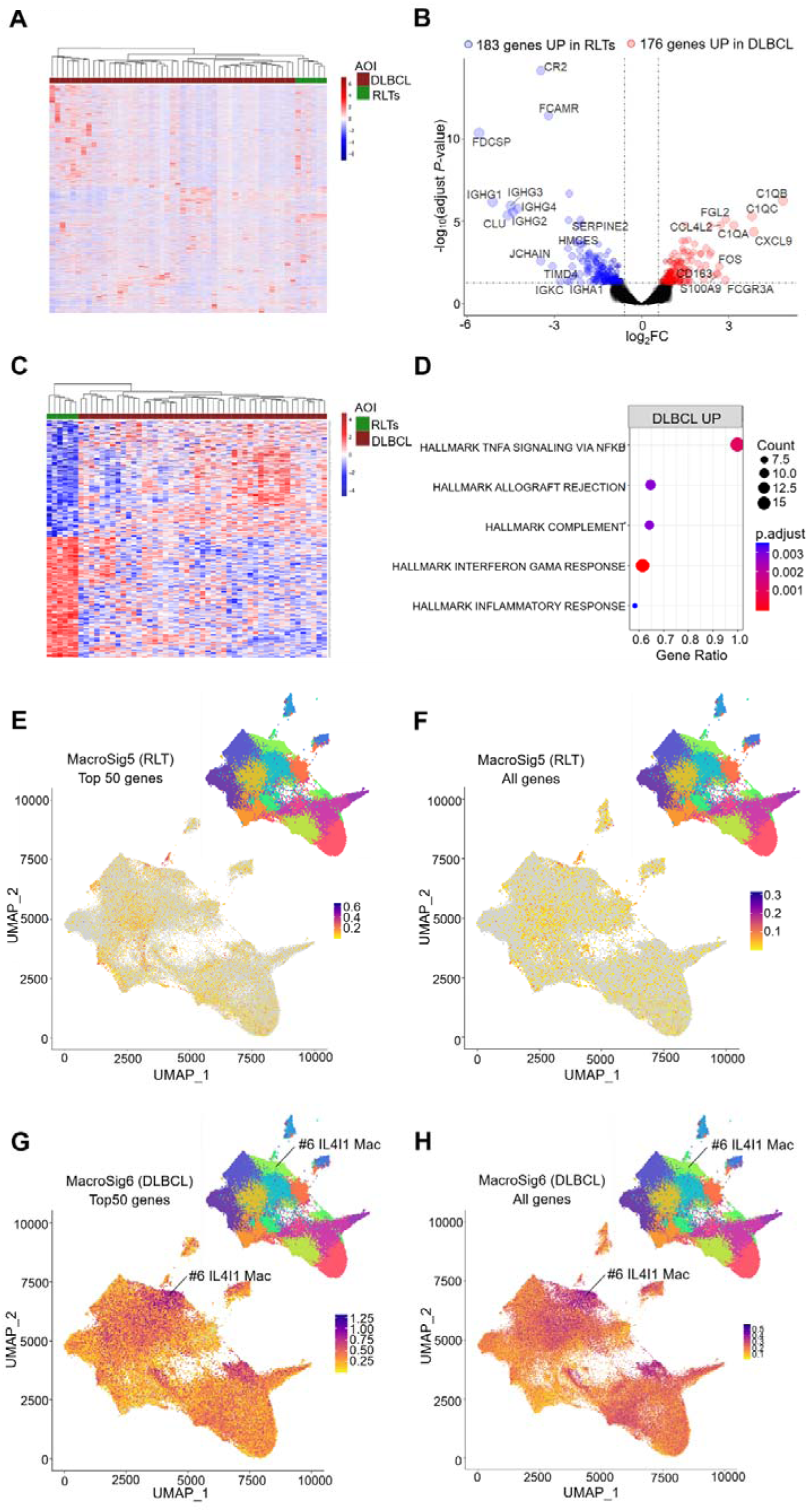
Distinct transcriptomic profiles of macrophages in RLTs and DLBCL. **(A)** Unsupervised clustering analysis was performed on transcripts from CD68+ masks between GC and DLBCL. **(B-C)** The volcano plot and heatmap showed the macrophage DEGs between GC and DLBCL based on adjusted *P*-value (p<0.05, |log_2_-FC|≥0.58). **(D)** GSEA was performed on the significant macrophage DEGs between GC and DLBCL (p<0.05, |log_2_-FC|≥0.58). **(E-H)** MacroSig5 (RLT) and MacroSig6 (DLBCL) were projected into MoMac-VERSE, respectively.

### MacroSigs from relapsed DLBCL patients map to IL1B monocytes

An unsupervised clustering analysis of genes obtained from the CD68+ mask in DLBCL patients revealed substantial inter-tumor heterogeneity (ward.D2 aggregation method). There were no clear clusters corresponding to different diagnostic sites, international prognostic index (IPI) scores, and cell-of-origin (COO) subtypes (Figure 4A). We then compared macrophage DEGs between two groups of cases with different clinical outcomes to R-CHOP (relapse (*N* = 16) vs no relapse (*N* = 29) with at least 3 years of follow up). While numerous genes were differentially expressed between both groups, there were no single DEGs with p < 0.05 after *P*-value correction (Figure 4B-C, Moderated *t*-test). The top differential genes (|log_2_ FC| > 0.58) were used to define MacroSig7 (No relapse) and MacroSig8 (Relapse). A GSEA of the top differential genes suggests that TNF-α, hypoxia, and inflammatory response signaling pathways were significantly upregulated in relapsed patients (Figure 4D, *P* < 0.005, Fisher’s exact test). The genes significantly involved in the TNF-α signaling pathway are displayed in Figure 4E.

**Figure 4.**
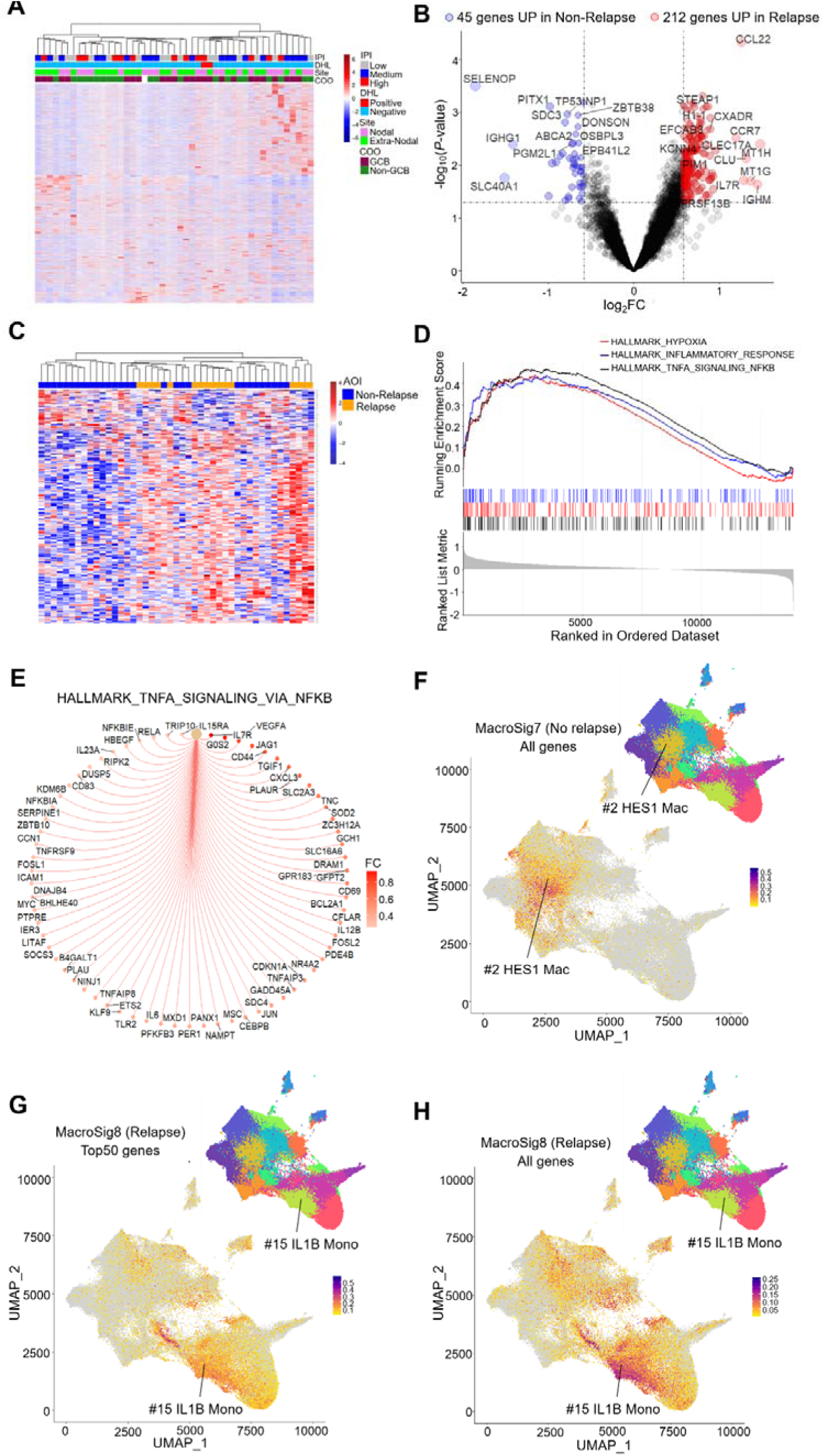
MacroSigs from relapsed DLBCL patients map to IL1B monocytes. **(A)** Unsupervised clustering analysis was performed on transcripts from CD68+ masks in DLBCL patients. **(B-C)** The volcano plot and heatmap showed the macrophage DEGs between DLBCL patients with or without relapse based on *P*-value (p<0.05, |log_2_-FC|≥0.58). **(D)** GSEA was performed on the all of macrophage DEGs between DLBCL patients with or without relapse. The top three enriched signaling pathway were displayed. **(E)** Gene-concept network plot displayed all genes that are involved in TNF-α signaling pathway**. (F-H)** MacroSig7 (No relapse) and MacroSig8 (Relapse) were projected into MoMac-VERSE, respectively. international prognostic index, IPI; double hit lymphoma, DHL; cell of origin, COO; germinal center B-cell like, GCB.

Upon projection of the MacroSig7 (No relapse) and MacroSig8 (Relapse) onto MoMac-VERSE, the MacroSig7 (No relapse) was distinctly located within the HES1/FOLR2 macrophage population (Figure 4F) while the MacroSig8 (Relapse) predominantly overlapped with IL1B monocytes that have been demonstrated to have pro-inflammatory effects on the TME in various cancers (Figure 4G-H) (33, 34). These results indicate that there are differences between TAM subclusters in the DLBCL TME in patients who undergo relapse; whether proinflammatory IL1B monocytes may be a contributory factor to DLBCL relapse will represent an important area for additional mechanistic research.

### Spatially-derived MacroSigs associate with COO and genetic DLBCL subclassifications

We next sought to understand the relationship of our spatially-derived MacroSigs to established DLBCL genetic and molecular subtypes, using eight publicly available DLBCL transcriptomic datasets. We found that across all eight datasets, the MacroSig1 (GC) was enriched in the germinal center B-cell like (GCB) DLBCL (Figure 5A-B, *P* < 0.05, Fisher’s exact test, 8/8 datasets), while MacroSig2 (IF) was enriched in unclassified (UNC) DLBCL (*P* < 0.005, Fisher’s exact test, 6/8 datasets), and MacroSig6 (DLBCL) in activated B-cell (ABC) subtype DLBCL (*P* < 0.01, Fisher’s exact test, 7/8 datasets,). The robust association (All *P*-values < 0.05) of these spatially derived MacroSigs to the B-cell gene expression profiling-based DLBCL COO subclassification highlights a diversity of molecular states in lymphoma-associated macrophages that complements the functional differentiation of the B-cell clones. MacroSig7 (No relapse) and MacroSig8 (Relapse) also segregated into GCB and ABC subtypes respectively (Figure 5C, *P* < 0.0001, Fisher’s exact test, 8/8 datasets), while the MacroSig3 (LZ) and MacroSig4 (DZ) did not clearly enrich any COO category (Figure 5D).

**Figure 5.**
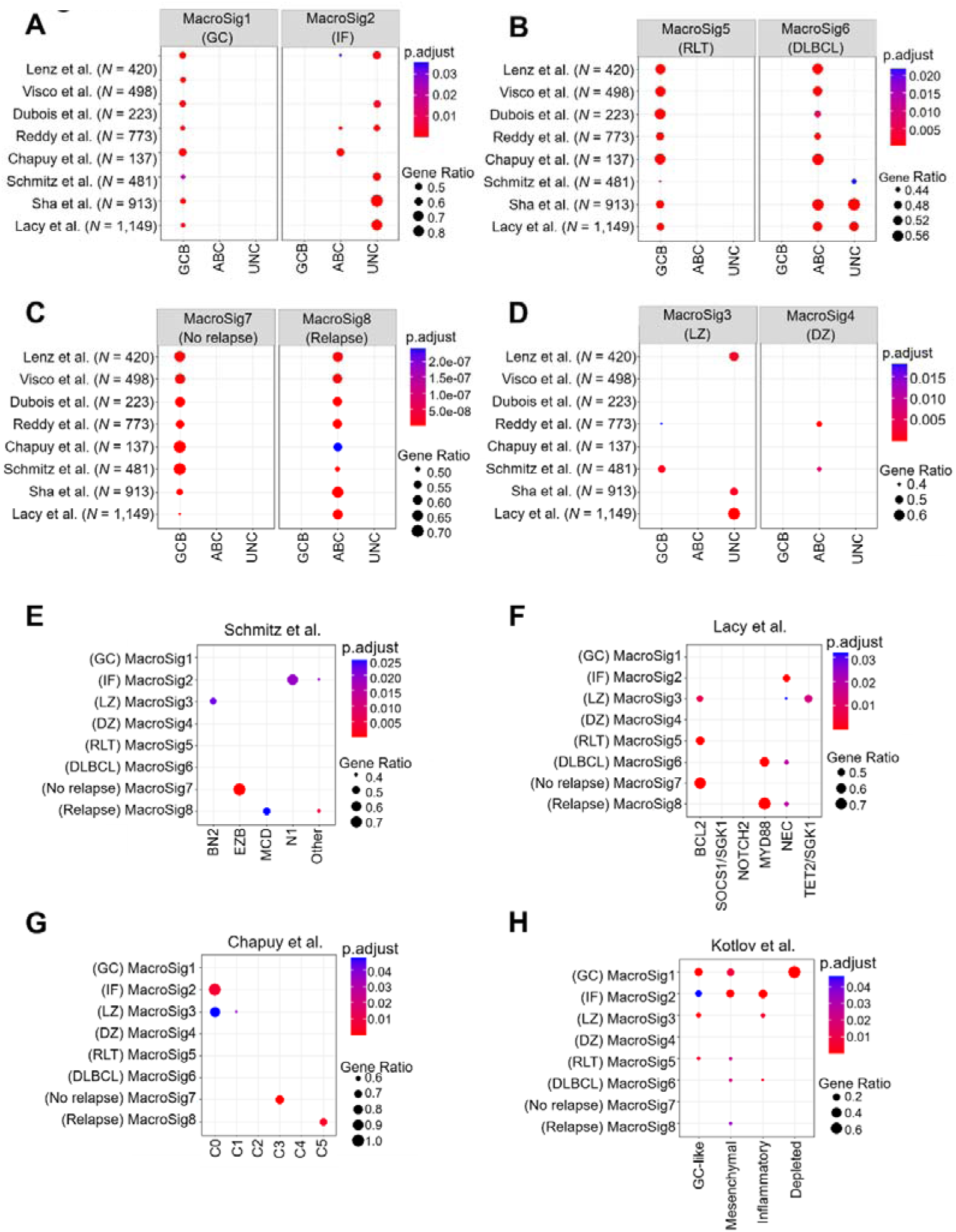
Spatially-derived MacroSigs associated with COO and genetic DLBCL subclassifications. **(A)** MacroSig1 (GC) and MacroSig2 (IF) were enriched in DLBCL COO classifications in eight publicly available datasets. **(B)** MacroSig5 (RLT) and MacroSig6 (DLBCL) were enriched in DLBCL COO classifications in eight publicly available datasets. **(C)** MacroSig7 (No relapse) and MacroSig8 (Relapse) were enriched in GCB and ABC DLBCL, respectively. **(D)** MacroSig3 (LZ) and MacroSig4 (DZ) were partially enriched in DLBCL COO classifications. Enrichment analysis of the eight MacroSigs was performed on the genetic and molecular subtypes of DLBCL in 3 distinct datasets of Schmitz et al **(E)**, Lacy et al **(F)** and Chaupy et al **(G). (H)** Enrichment analysis of the eight MacroSigs was performed on DLBCL microenvironment categories generated by Kotlov et al. activated B-cell like, ABC.

We then examined the relationship between these MacroSigs and genetic subtypes in 3 distinct datasets. In Schmitz et al (35), MacroSig3 (LZ) and MacroSig7 (No relapse) were respectively enriched in BN2 and EZB subtypes that are associated with more favorable progression-free survival (PFS) and OS (Figure 5E, *P* < 0.005, Fisher’s exact test). In contrast, MacroSig2 (IF) and MacroSig8 (Relapse) were respectively enriched in N1 and MCD subtypes that are related to poorer PFS and OS (*P* < 0.01, Fisher’s exact test). Concordantly, in the dataset from Lacy et al (36), MacroSig3 (LZ), MacroSig5 (RLT), and MacroSig7 (No relapse) were mostly enriched in BCL2 subtypes that showed good OS (Figure 5F, *P* < 0.05, Fisher’s exact test). MacroSig6 (DLBCL) and MacroSig8 (Relapse) were mainly enriched in MYD88 subtypes that showed poor OS (*P* < 0.0005, Fisher’s exact test). In Chapuy et al (37), MacroSig2 (IF) and MacroSig3 (LZ) were enriched in the C0 subtype (Figure 5G, *P* = 0.006 and *P* < 0.05, respectively, Fisher’s exact test). MacroSig7 (No relapse) and MacroSig8 (Relapse) were enriched in the C3 and C5 subtypes, respectively (*P* < 0.0001 and *P* < 0.005, respectively, Fisher’s exact test). Beyond genetic classifications, we also evaluated the relationship of our MacroSigs with DLBCL TME categories proposed by Kotlov et al (5). No clear enrichment of our MacroSigs with specific DLBCL TME categories emerged (Figure 5H, Fisher’s exact test). Overall, these results show a consistent association of our spatial MacroSigs to specific genetic subgroups, suggesting that MacroSigs may provide a novel microenvironment subcategorization that may complement current genetic DLBCL classifications.

### Spatially-derived MacroSigs stratify for patient survival in DLBCL datasets

Finally, we aimed to assess the prognostic significance of these newly defined spatially-derived MacroSigs by exploiting the above-mentioned eight DLBCL datasets. Across most datasets, no prognostic significance of MacroSig1 (GC)- and MacroSig2 (IF) was observed (data not shown). In contrast, survival analysis showed that cases with the MacroSig6 (DLBCL) had shorter OS than that of cases with the MacroSig5 (RLT) (Figure 6A, *P* < 0.05, 5/8 datasets; hazard ratios (HR) and 95% confidence intervals (CI) for each dataset in Supplementary Table S4), as demonstrated in the Kaplan-Meier plots across five datasets (Figure 6B-F, *P* < 0.05, Log-rank test). Similarly, across five datasets, patients with MacroSig8 (Relapse) had significantly shorter OS compared to those with MacroSig7 (No relapse) (Figure 6G-L, *P* < 0.05, 5/8 datasets, Log-rank test; HR and 95% CI refer to Supplementary Table S4).

**Figure 6.**
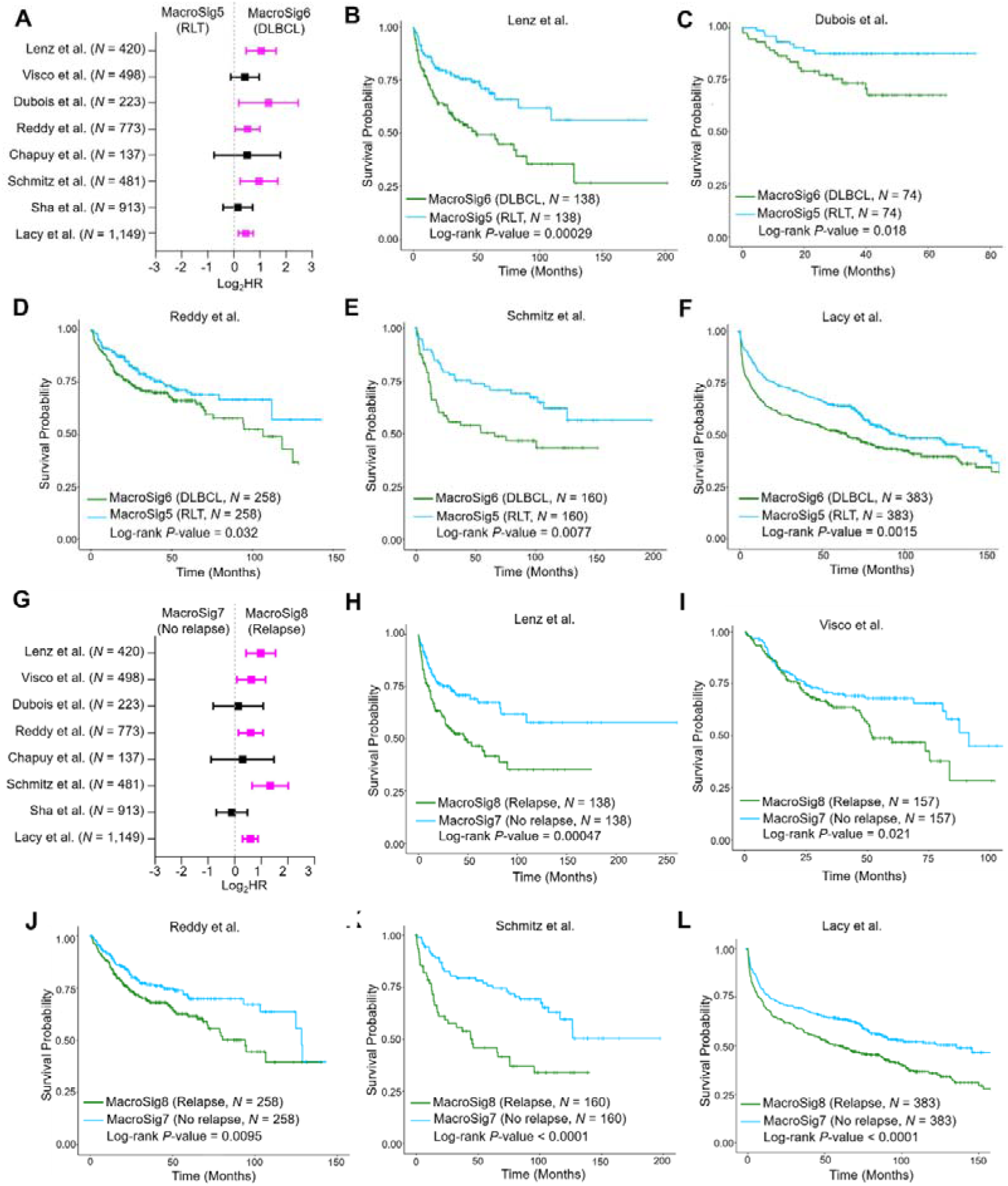
Spatially-derived MacroSig5-8 stratify for patient survival in DLBCL datasets. **(A)** Univariate Cox proportional hazards model analysis for MacroSig5 (RLT) and MacroSig6 (DLBCL) in eight publicly available DLBCL datasets. **(B-F)** Kaplan–Meier analyses showed that MacroSig6 (DLBCL) was associated with poor OS in DLBCL patients across five distinct DLBCL datasets. **(G)** Univariate Cox proportional hazards model analysis for MacroSig7 (No relapse) and MacroSig8 (Relapse) in eight publicly available DLBCL datasets. **(H-L)** Kaplan–Meier analyses showed that MacroSig8 (Relapse) was associated with poor OS in DLBCL patients across five distinct DLBCL datasets. overall survival, OS.

We observed that DLBCL patients with MacroSig4 (DZ) had significantly worse OS compared to those with MacroSig3 (LZ) across all eight DLBCL datasets (Figure 7A-I, *P* < 0.05, 8/8 datasets, Log-rank test; HR and 95% CI refer to Supplementary Table S4). This association was also validated using a multivariate analysis (Supplementary Table S5, *P* < 0.05, 5/8 datasets (IPI sore is not available in 2 datasets) adjusted for the factors of the IPI score across most datasets. In contrast to B-cell-based LZ and DZ signatures, LZ- and DZ-MacroSigs (MacroSig3-4) were much better predictors of patient prognosis (Supplementary Figure S4A-I, *P* < 0.05, 4/8 datasets, Log-rank test). Only few genes were repeated across B-cell derived and macrophage-derived LZ- and DZ-signatures (MacroSig3-4) (Figure 7J), which demonstrated that the prognostic value of macrophage transcriptional heterogeneity within the LZ and DZ is independent of B cell transcriptome. Likewise, although both MacroSig4 (DZ), MacroSig6 (DLBCL), and MacroSig8 (Relapse) are predictive of poor prognosis in DLBCL patients, few genes overlap across these signatures (Figure 7K and Supplementary Table S3). These results shows that the MacroSigs we experimentally derived from DSP are distinct and confer prognostic significance. This new information in the complex landscape of DLBCL may contribute to improved delineation of patients’ prognosis and may provide impetus for development of novel macrophage-centered treatments.

**Figure 7.**
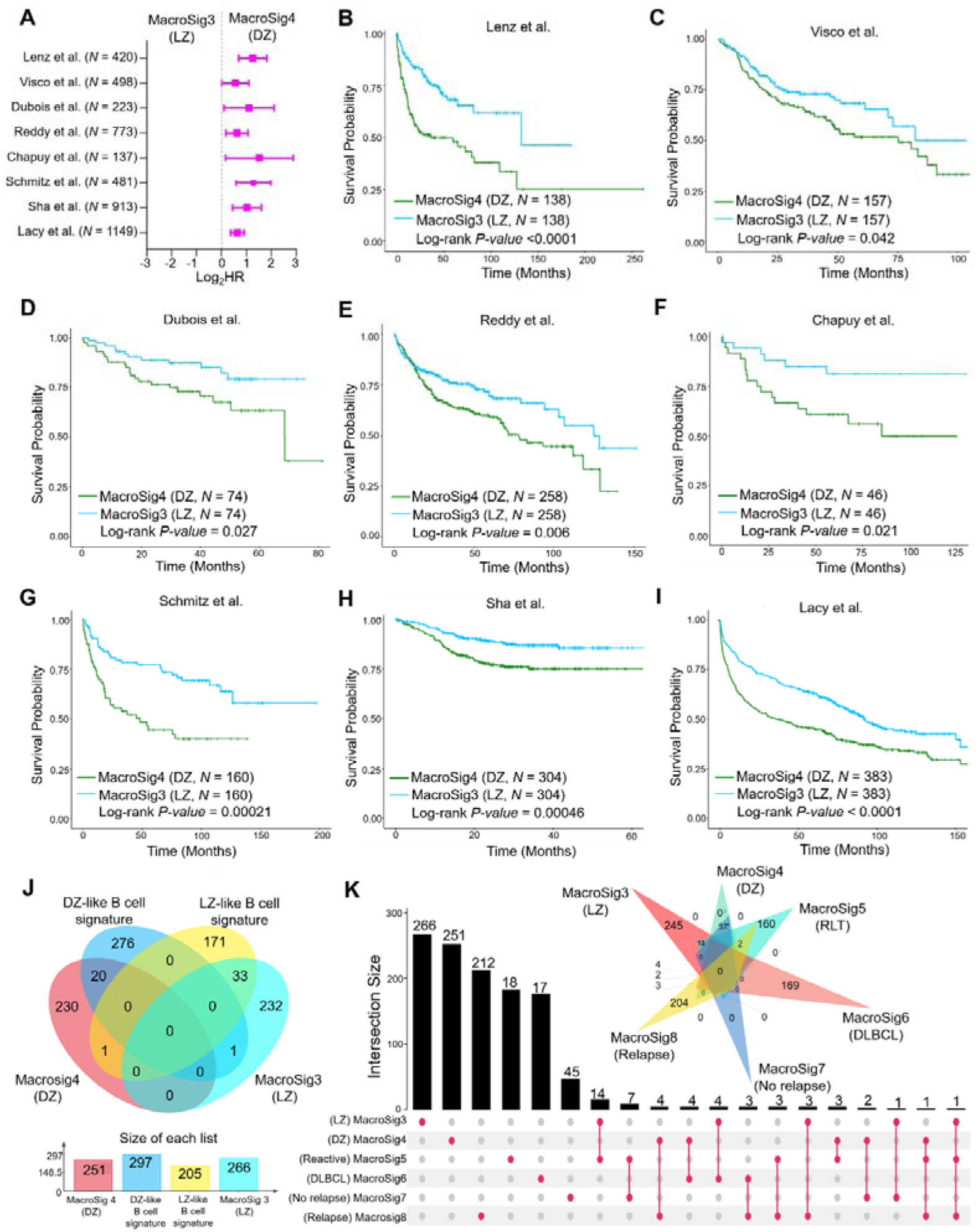
Spatially-derived MacroSig3-4 stratify for patient survival in DLBCL datasets. **(A)** Univariate Cox proportional hazards model analysis for MacroSig3 (LZ) and MacroSig4 (DZ) in eight publicly available DLBCL datasets. **(B-I)** Kaplan–Meier analyses showed that MacroSig4 (DZ) was associated with poor OS in DLBCL patients across eight distinct DLBCL datasets. **(J)** Venn diagram displayed the overlapping genes in LZ-, DZ-like B-cell signatures, and MacroSig3-4 (LZ and DZ). **(K)** UpSet and Venn diagram showed the overlapping genes in our six MacroSigs.

## Discussion

Using DSP, we defined eight macrophage transcriptomic signatures in reactive and malignant lymphoid tissue (termed MacroSig1-8; corresponding to biological/clinical categories listed in Table 1) and described their associations with known features of macrophage/ DLBCL biology and clinical outcome (Graphical Abstract). Macrophages are highly plastic, acquiring diverse phenotypes and functions that enable their role in a variety of physiological or pathological settings (38). We demonstrate for the first time the spatial transcriptional diversity of macrophages within different regions of RLTs and between macrophages populating RLTs and DLBCL. In RLTs, we show that GC-associated macrophages exhibit a transcriptional profile clearly distinct from that of macrophages in IF regions. The development of concise macrophage gene signatures that are representative of their molecular and functional diversity, but which are also clinically applicable, is an area of ongoing research (39). Importantly these two subsets (GC/IF) do not map to any previously defined macrophage groups based on scRNA-seq (27), highlighting a potential value of spatially resolved profiling for other tissue contexts. Among the hallmark genes of GC-macrophages was the matricellular protein SPARC. We and others have demonstrated the involvement of SPARC in regulating the inflammatory profile of innate immune cells in lymphomagenesis and in the organization of DLBCL TME (40, 41). Macrophage-associated SPARC expression has been reported as a stromal marker associated with a favorable outcome in independent case series (11, 42). Consistently, we also see better survival in patients with the GC-MacroSig.

**Table 1.**
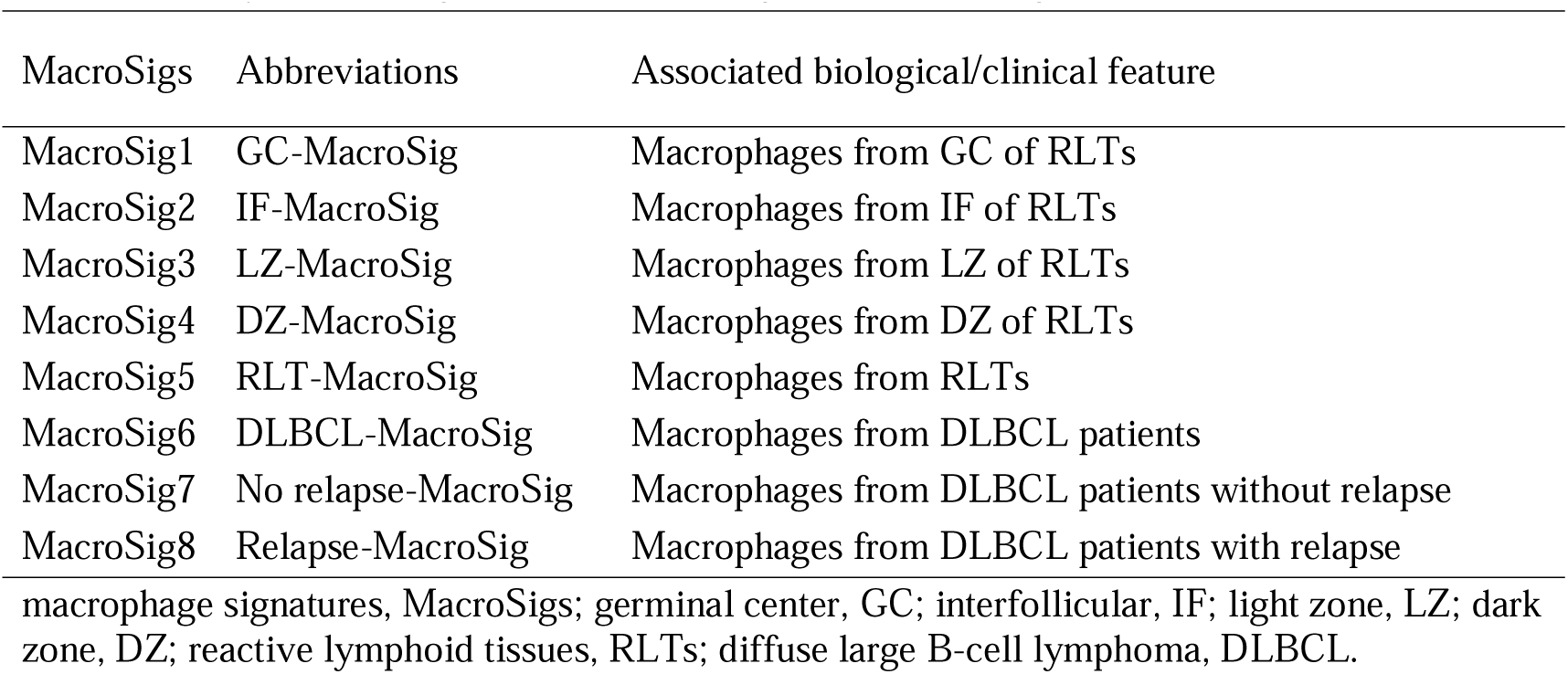
Newly-defined eight distinct MacroSigs based on biological/ clinical characteristics

| MacroSigs | Abbreviations | Associated biological/clinical feature |
| --- | --- | --- |
| MacroSig1 | GC-MacroSig | Macrophages from GC of RLTs |
| MacroSig2 | IF-MacroSig | Macrophages from IF of RLTs |
| MacroSig3 | LZ-MacroSig | Macrophages from LZ of RLTs |
| MacroSig4 | DZ-MacroSig | Macrophages from DZ of RLTs |
| MacroSig5 | RLT-MacroSig | Macrophages from RLTs |
| MacroSig6 | DLBCL-MacroSig | Macrophages from DLBCL patients |
| MacroSig7 | No relapse-MacroSig | Macrophages from DLBCL patients without relapse |
| MacroSig8 | Relapse-MacroSig | Macrophages from DLBCL patients with relapse |

macrophage signatures, MacroSigs; germinal center, GC; interfollicular, IF; light zone, LZ; dark zone, DZ; reactive lymphoid tissues, RLTs; diffuse large B-cell lymphoma, DLBCL.

Conversely, we identify novel MacroSigs that are prognostic for poor clinical outcomes in multiple independent DLBCL datasets. MacroSig6 (DLBCL) was also negatively prognostic, and mostly corresponded to the IL4I1 macrophage population. IL4I1 exerts an immunosuppressive effect by dampening the antitumor CD8+ T-cell response, resulting in tumor immune evasion (32, 43). Thus, blockade of IL4I1 activity in macrophages may facilitate the remodeling of specific anti-tumor immune responses (44). Complement pattern recognition components (C1QA, C1QB, and C1QC, etc) were also significantly upregulated in DLBCL macrophages compared to GC-macrophages. While further validation is required for C1Q+ macrophages in DLBCL, C1Q is a promising prognostic marker and therapeutic target in other hematological malignancies. High C1Q expression in macrophages-like leukemia cells has been associated with extramedullary infiltration and poor prognosis in acute myeloid leukemia (45). The other negatively prognostic signature in our study was MacroSig8 (Relapsed-DLBCL), which maps to IL1B pro-inflammatory monocytes with pro-tumorigenic effects (46). Gevokizumab, a monoclonal antibody targeting IL1B, is testing in clinical trials for its efficacy in treatment of metastatic cancers (NCT03798626), and could represent a potential therapeutic strategy in combination with chemotherapy for tumors with high infiltration by this macrophage subtype. Using Gene Ontology Enrichment Analysis, we also observed many ion channels or divalent metal transporters overexpressed in these relapsed-DLBCL macrophages. Zinc and copper have been described in macrophage-mediated host defense and antimicrobial pathways (47–49), but how divalent metal transporters control TAMs functions in relation to chemotherapy warrants further investigation. It is possible that these ion channels could be a potential immunomodulatory mechanism in TAMs, supporting DLBCL progression during treatment, and offering a novel scenario for therapeutic targeting.

Another key finding from our study is the association of certain MacroSigs with established clinically-relevant DLBCL subclassifications (5, 41, 42, 50). Gene expression profiling identifies two prognostically-distinct clusters of DLBCL based on COO, but there remains a consistent cluster of “Unclassified” cases. We report a strong correlation of the signature of IF-macrophages (MacroSig2) with this Unclassified cluster, along with clear correlations of MacroSig1 (GC) to the GCB DLBCL cluster and MacroSig6 (DLBCL) to the ABC cluster. These results suggest that DLBCL gene expression signatures may be entwined with the nature of underlying macrophage infiltrates, and the role of IF-like macrophages in potentially determining the overall gene-expression status of the previously unclassified COO subcluster will be an interesting avenue of research.

Molecular high-grade (MHG) (51)/ double-hit gene expression signature (DHITsig) (52) is another salient molecular subgroup of interest in DLBCL. This subgroup carries a uniformly poor prognosis and is thought to be reflective of GC-DZ biology (23, 53), and was renamed the DZ signature (53). It is intriguing that the strongest prognostic signature in our study was also of DZ-cells, but that of macrophages and not B-cells. Importantly, we found that genes comprising MacroSig4 (DZ), collected from CD68+ DZ macrophages, are distinct from published B-cell DZ signatures and from those collected in our own experiments from CD20+ DZ B-cells. Notably, both MacroSig4 (DZ) and B-cell DZ signatures were highly prognostic in multiple DLBCL datasets, with higher effect sizes and more consistent statistical significance for the MacroSig4 (DZ). None of genes constituting MacroSig4 (DZ) were noted to be highly expressed within CD20+ cells in DLBCL in our dataset, indicating that the signature is likely to confer its prognostic significance because of macrophage infiltration and not aberrant expression of these genes in tumor cells. Pathway enrichment analysis indicated that the cell proliferation/cycle-associated pathways were activated in DZ macrophages. A recent transcriptomic analysis of the circulating MDMs identified a proliferative macrophage subcluster which influence MDMs-mediated inflammation and regeneration (54). It is tempting to speculate a similar biology exists in these proliferative DZ macrophages, the molecular (proteomic/secretomic) characteristics of DZ macrophages of which require further investigation.

A potential limitation of DSP is its reliance on average values across a group of cells defined by a cellular “mask”, and its resolution which likely extends beyond the size of a single cell. Nonetheless, we have demonstrated that there is minimal overlap of gene sets between the CD68+ and CD20+ masks profiled regions, suggesting that the technical spill-over of transcripts between the CD68 and CD20 photocleaved DSP regions is not a driver of the observed variability among different macrophage spatial profiles. While our study has provided novel insight into macrophage transcriptomic subtypes in RLTs and DLBCL tissues, the active scavenging function of macrophages may contribute to non-macrophage transcript contamination (55), thus posing a further limitation of DSP analyses. Further refinements to spatial transcriptomics at single-cell resolution will provide more detailed dissection of intrinsic vs scavenged transcripts, and also allow direct evaluation of spatial interactions between different cell types, contributing to a deeper understanding of the actual relationships between macrophage subtypes and other TME components.

In summary, through the use of DSP, we demonstrate the first spatially resolved transcriptomic characterization of macrophages in RLTs and DLBCL, showing diverse characteristics of macrophage landscapes in different spatial localizations. Spatially-derived MacroSigs of lymphoid tissue can complement existing genetic and molecular DLBCL subclassifications, contributing to our understanding of DLBCL TME and the ever-evolving classification of this heterogenous disease, and are also robust in predicting prognosis across numerous distinct DLBCL datasets. These data provide a framework to further evaluate the biological and clinical relevance of macrophage subtypes in lymphoid biology and disease.

## Methods

### DSP study population

DLBCL samples in a tissue microarray (TMA) format were derived from pre-treatment biopsies of 47 patients treated with 6x R-CHOP between 2010 and 2017 at the National University Hospital in Singapore (Group 1). The characteristics of these patients are summarized in Supplementary Table S1, with a median follow-up time of six years. Group 2 (non-malignant control samples) comprises a TMA of 12 RLTs (tonsil) samples and 5 individual RLT whole-slide samples, obtained from patients with tonsillectomies at the National University Hospital for non-cancer indications. All tissues were formalin-fixed and paraffin-embedded at the time of resection, and stored as paraffin blocks.

### GeoMx® DSP WTA assay

Spatial transcriptomics of RLTs and DLBCL tissues was conducted using the GeoMx WTA kit (NanoString, Seattle, Washington, USA), according to manufacturer’s instructions. Antibodies used, staining and sample collection, as well as library preparation and sequencing methods are detailed in Supplementary Methods.

### Datasets

DLBCL expression array datasets of Lenz et al (*N* = 420) (42), Visco et al (*N* = 498) (56), and Dubois (*N* = 223) (57) were obtained from Gene Expression Omnibus, while DLBCL RNA-seq datasets of Reddy et al (*N* = 773) (58), Chapuy et al (*N* = 137) (37), Schmitz et al (*N* = 481) (35), and Sha et al (*N* = 913) (51), Lacy et al (*N* = 1,149) (36) were obtained from the database of Genotypes and Phenotypes. Single-cell RNA sequencing (scRNA-seq) datasets of RLTs were obtained from King et al (*N* = 3) (59) and Stewart (*N* = 7) (60), and scRNA-seq datasets of DLBCL from Steen et al (*N* = 7) (61) and Roider et al (*N* = 3) (62).

### Data processing and analyses

Quality control and normalization of the DSP data was processed using GeoMx DSP software v2.4.0.421. The ward.D2 aggregation method was used for clustering analysis across regions of interest. Supplementary Methods contain further details on raw data processing and subsequent analyses which include differential expression analyses, gene set enrichment analysis (GSEA), enrichment analysis, and scRNA-seq data analysis. Statistical analyses were conducted on R statistical software (v 4.1.2) (http://www.R-project.org). All *P*-values were two-sided and considered statistically significant when <0.05. All analyses were REMARK compliant and the checklist is provided in Supplementary Data.

### Derivation of spatial MacroSigs and survival analysis

We compared the differential genes of macrophages in four paired analyses: GC versus IF, LZ versus DZ, RLT versus DLBCL, and Non-relapse-DLBCL versus Relapse-DLBCL. Macrophage “signatures” were then derived based on genes enriched in a particular group, fulfilling one of two criteria: 1) adjusted *P*-values < 0.05 applying the Benjamini-Hochberg correction on moderated p-values (Limma); 2) absolute log_2_ fold change (|log_2_FC| > 0.58). Eight MacroSigs were derived from the upregulated/downregulated genes corresponding to the four groups. To evaluate the prognostic power of our MacroSigs, we applied each MacroSig group (upregulated/downregulated genes) into each of the eight aforementioned DLBCL datasets using a quantile strategy. Patients were divided into three groups: 1) those with high expression of downregulated genes; 2) with an intermediate gene signature expression; 3) with high expression of upregulated genes. Overall survival (OS) of patients in extreme groups (i.e., group 1 and group 3) were compared using Log-rank test and Cox proportional hazards model. More details are described in the Supplementary Methods.

### Study approval

All biopsy samples were obtained from the Department of Pathology, National University Hospital, in accordance with the ethical guidelines of the domain specific review board (DSRB) approved protocol 2015/00176.

## Author contributions

ML, CT and ADJ conceived and designed the study. FSY, YFP, JT, RC, SDM, LP, EC, JL, YLC, and SBN prepared the specimens. ML performed the experiments. ML, GB, KM, SS, PJ, FG, CT, and ADJ performed data analysis. MMH, JT, RC, SDM, LP, EC, JL, and YLC participated in the collection of clinical data. ML, RXL, PJ, CT and ADJ wrote the manuscript. KGC offered some suggestions of manuscript wring. All authors edited and approval the final version of the manuscript.

## Disclosures of Conflicts of Interest

ADJ has received consultancy fees from Roche, Gilead, Turbine Ltd, AstraZeneca, Antengene, Janssen, MSD and IQVIA, and research funding from Janssen and AstraZeneca. The other co-authors have no relevant conflicts of interest to declare.

## Supporting information

The supplementary data will be used for the link to the file on the preprint site.

## Data Availability

All data produced in the present study are available upon reasonable request to the authors.

## Acknowledgements

ML was supported by the China Scholarship Council (202006940018). ADJ was supported by the Singapore Ministry of Health’s National Medical Research Council Clinician Scientist Award (MOH-000715-00). Work in ADJ’s laboratory is funded by a core grant from the Cancer Science Institute of Singapore, National University of Singapore through the National Research Foundation Singapore and the Singapore Ministry of Education under its Research Centres of Excellence initiative. CT was supported by the Italian Foundation for Cancer Research (AIRC) Investigator Grant IG ID.22145; 5x1000 Grant ID.22759.

## Notes

### Competing Interest Statement

The authors have declared no competing interest.

### Funding Statement

ML was supported by the China Scholarship Council (202006940018). ADJ was supported by the Singapore Ministry of Health National Medical Research Council Clinician Scientist Award (MOH-000715-00). The study is funded by a core grant from the Cancer Science Institute of Singapore, National University of Singapore through the National Research Foundation Singapore and the Singapore Ministry of Education under its Research Centres of Excellence initiative. CT was supported by the Italian Foundation for Cancer Research (AIRC) Investigator Grant IG ID.22145; 5x1000 Grant ID.22759.

