## Supplementary material for "Spatially-Resolved Transcriptomics Define Clinically Relevant Subsets of Macrophages in Diffuse Large B-cell Lymphoma": The supplementary data will be used for the link to the file on the preprint site.

**Supplementary Appendix: Spatially Resolved Transcriptional profiling of Macrophages in Diffuse Large B-cell Lymphoma**

Patients	[3](#_Toc113395705)

GeoMx® DSP sample preparation on Bond Max	[3](#_Toc113395705)

In-situ hybridization	[3](#_Toc113395706)

GeoMx® DSP sample collection	[3](#_Toc113395706)

Library preparation and sequencing	[4](#_Toc113395706)

Quality control and normalization of DSP data	5

Differential expression analyses	[5](#_Toc113395707)

Gene set enrichment analysis	[5](#_Toc113395708)

Correlation analysis	[5](#_Toc113395709)

Survival analysis	[6](#_Toc113395710)

Single-cell sequencing data analysis	[7](#_Toc113395711)

[Supplementary Tables](#_Toc113395714)	8

### Investigators

**Authors:** Min Liu^1,2,3^; Giorgio Bertolazzi^4^; Kevin Mulder^5,6,7^; Shruti Sridhar^1^; Rui Xue Lee^1^; Patrick Jaynes^1^; Michal Marek Hoppe^1^; Shuangyi Fan^8^; Yanfen Peng^1^; Jocelyn Thng^1^; Reiya Chua^9^; Sanjay De Mel^9,10^, Limei Poon^9,10^, Esther Chan^9,10^, Joanne Lee^9,10^ Susan Swee-Shan Hue^8,10^, Siok-Bian Ng^1,8,10^; K George Chandy^11^, Florent Ginhoux^5,6,7^; Yen Lin Chee^9,10^; Claudio Tripodo^4,12*^; Anand D. Jeyasekharan^1,9,10,13*^

**Affiliations:**

^1^Cancer Science Institute of Singapore, National University of Singapore, Singapore

^2^Department of Radiation Oncology, Chongqing University Cancer Hospital, Chongqing, P. R. China

^3^Department of Immunology, Tianjin Medical University Cancer Institute and Hospital, P. R. China

^4^Tumor Immunology Unit, University of Palermo, Palermo, Italy

^5^Singapore Immunology Network, Agency for Science, Technology and Research, Singapore

^6^Institut National de la Santé Et de la Recherche Medicale (INSERM) U1015, Equipe Labellisée—Ligue Nationale contre le Cancer, 94800 Villejuif, France

^7^Université Paris-Saclay, Gustave Roussy, Villejuif, France

^8^Department of Pathology, Yong Loo Lin School of Medicine, National University of Singapore, Singapore

^9^Department of Haematology-Oncology, National University Health System, Singapore

^10^NUS Centre for Cancer Research, Yong Loo Lin School of Medicine, National University of Singapore, Singapore

^11^Lee Kong Chian School of Medicine, Nanyang Technological University Singapore, Experimental Medicine Building, 59 Nanyang Drive, 636921, Singapore

^12^IFOM ETS - The AIRC Institute of Molecular Oncology, 20139 Milan, Italy

^13^Department of Medicine, Yong Loo Lin School of Medicine, National University of Singapore, Singapore

### Supplementary Methods

**Patients**

47 patients were diagnosed with primary diffuse large B cell lymphoma (DLBCL). Patients were randomly selected. Clinical endpoint was overall survival after six cycles of immunochemotherapy combination of rituximab with cyclophosphamide, doxorubicin, vincristine, and prednisolone.

**GeoMx® DSP sample preparation on Bond Max**

The formalin-fixed paraffin-embedded sections were freshly cut (5µm thick) and placed on Poly-L-lysine-coated microscope glass slides (Leica Biosystems, Germany). The slides were baked at 60°C for 1 h and loaded, with covertiles, into the slide tray on Bond Max (Leica Biosystems) to run deparaffinization, rehydration, antigen retrieval (ER2 solution (Leica Biosystems) at 100 °C for 20 min), RNA digestion (Proteinase K 1μg/ml for 15 min) and post-fixation (10% neutral buffered formalin (NBF, Sigma-Aldrich, St. Louis, Missouri, USA) for 5 min, NBF stop buffer for 5 min twice). The NBF stop buffer was prepared using Tris base (Promega, Madison, Wisconsin, USA) and Glycine (Thermo Fisher Scientific, Waltham, Massachusetts, USA) in DEPC-treated water. Once the run has finished, the covertiles were removed and the slides were soaked in PBS for subsequent hybridization.

**In-situ hybridization**

An overnight in-situ hybridization was performed with GeoMx® Human NGS whole transcriptome atlas (NanoString, Seattle, Washington, USA) that contained probes for 18,000+ protein-coding genes. After which, the slides were washed twice with equal parts of 4X SSC (Thermo Fisher Scientific) and 100% formamide (Sigma-Aldrich) at 37 °C for 25 min to remove off-target probes.

**GeoMx****®** **DSP sample collection**

The slides were incubated with blocking Buffer W (200 μL/slide, NanoString) for 30 min in the humidity chamber at room temperature after hybridization. For Group 1, slides of DLBCL TMA were stained with macrophage marker CD68 (sc-20060 AF594, Santa Cruz biotechnology, Texas, USA), T-cell marker CD3 (Dako, A0452, California, USA), B-cell marker CD20 (Novus Biologicals, NBP2-47840 AF647, Colorado, USA) and the nuclear stain SYTO 13. For Group 2 (Figure 1B), the slide of RLTs TMA was visualized with CD68, the follicular dendritic cell marker nerve growth factor receptor (NGFR, Ab52987, Abcam, Cambridge, UK) and SYTO 13. Individual RLT sections of Group 2 (Supplementary Figure S1B) were stained with similar markers as Group 1. Additionally, corresponding serial sections were stained with NGFR to identify light zone (LZ) and dark zone (DZ) regions. After immunofluorescent staining, the slides were visualized using the GeoMx® DSP instrument to select regions of interest (ROIs). To acquire representative regions, ROI selection was performed by an expert pathologist. The pathologist was blinded to the group assignment during the ROIs selection. Based on fluorescent staining, each ROI was segmented into corresponding areas of interest (AOIs) - CD68+ regions, CD3+ regions, and CD20+ regions. Subsequently, each AOI was exposed to UV light and photocleaved oligos were aspirated from the solution into the wells of a collection plate (Figure 1A).

**Library preparation and sequencing**

Collected photocleaved oligos were PCR amplified with the corresponding GeoMx® Seq Code Primer Plate and Master Mix (NanoString). PCR products were pooled and cleaned with AMPure XP beads (Agilent, Santa Clara, California, USA) twice to obtain the libraries. The quality and concentration of libraries were assessed using a high sensitivity DNA Kit (Agilent) and Bioanalyzer. Subsequently, libraries were sequenced on an illumina sequencing platform (Hiseq or NovaSeq) with standard workflow specifications (dual-indexing and paired-end reads (2 × 27 bp)).

**Quality control and normalization of DSP data**

Raw reads were trimmed, stitched, aligned, and deduplicated to transform digital counts data with unique targets. AOIs with fewer than 10,000 raw reads or sequencing saturation <50% were filtered out of the analysis. AOIs with less than 5% of all target genes (18,000+) and target genes that do not achieve the limit of quantitation were removed. The data was then processed with the Q3 normalization method for all the remaining targets. Q3 normalization divides the counts in one segment by the 3rd quartile value for that segment, then subsequently multiplies that value by the geometric mean of the 3rd quartile values of all segments. Q3 normalization rescales the gene expression data such that all segments have similar gene expression ranges. It reduces variance from segment size, segment cellularity, and other technical factors.

**Differential expression analyses**

Differential expression analyses were carried out using the limma package's moderated t-test.^1^ Upregulated/downregulated genes were selected by applying the Benjamini-Hochberg correction on the p-values (adjusted p-values < 0.05). In case of the absence of statistically significant genes, upregulated/downregulated genes were selected by applying a threshold on the absolute log_2_-fold-change (|log_2_FC| > 0.58).

**Gene set enrichment analysis (GSEA)**

The GSEA analyses were performed on R software (v 4.2.2). Hallmark gene sets were downloaded from Human Molecular Signature Database (MSigDB).^2^

**Correlation analysis**

The association between patient groups and clinical categories (i.e., cell-of-origin (COO) and genetic subtypes) have been evaluated through the Fisher exact test. The p-values have been adjusted for multiple comparisons.

**Survival analysis**

To evaluate the predictive power of our gene signatures from a clinical point of view, we have applied our signature on eight distinct diffuse large B-cell lymphoma (DLBCL) cohorts.^3-10^ We test each signature couple (upregulated/downregulated genes) using a quantile strategy to split each cohort into three groups. The patient ranking has been obtained by calculating the following score:

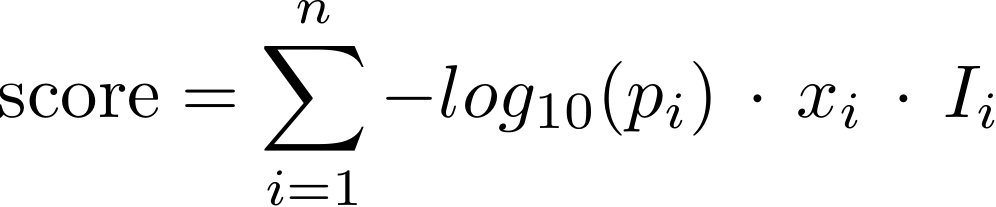

with,

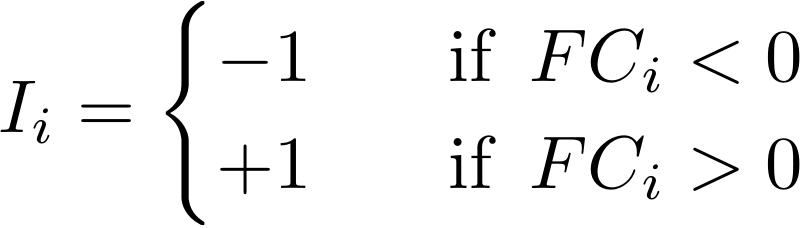

where *p_i_* and FC_i_ are the moderated t-test p-value and the FC of gene-*i* calculated running the differential expression analysis on the digital spatial profiling (DSP) data, *x_i_* is the expression of gene-*i* from the bulk RNA-seq data, and *n* is the number of genes of the gene signature. Using this score, each DLBCL cohort was divided into three groups according to the tertile values: 1) patients having high expression of downregulated genes - 2) patients having an intermediate gene signature expression - 3) patients having high expression of upregulated genes. The extreme groups (i.e., group 1 and group 3) have been compared in terms of overall survival (OS), COO, and genetic subtypes categories. Before calculating the log-rank test and fitting the Cox model, the cox.ph test was used to test the proportional hazard assumption. Kaplan-Meier method has been used to estimate the survival functions among groups, and the log-rank test has been used to test the differences in the OS between the selected groups. The Cox model has been fitted to estimate the relative risk of death between groups.

**Single-cell sequencing data analysis**

Seurat (v2.3.0)^11^ was used for the analysis of the single cell datasets. All functions were run with default parameters, unless specified otherwise. Low quality cells (<200 genes/cell, <3 cells/gene and >10% mitochondrial genes) were excluded. Harmony^12^ was used to integrate the single cell datasets and remove any batch effects. SingleR^13^ was used for annotation of clusters. Refinement and validation of annotation was conducted by projecting and evaluating a curated B cell, T cell and macrophage marker list.

For each macrophage signature (GC- and IF like, LZ- and DZ-like, GC- and DLBCL-like, Non-relapse- and relapse-like), the top 50 or all genes were taken to create a Module Score to analyze feature expression. This score was then projected onto the uniform manifold approximation and projection (UMAP) space named “Mo-MacVERSE”. The region that showed enrichment was further subsetted and analyzed.

### Supplementary Tables

| Supplementary Table S1. DLBCL patient characteristics | | | | |
| --- | --- | --- | --- | --- |
| Variables | | N | Percentage | |
| Age | |  |  | |
| ≤60y | | 21 | 44.7% | |
| ＞60y | | 26 | 55.3% | |
| Sex | |  |  | |
| Male | | 29 | 61.7% | |
| Female | | 18 | 38.3% | |
| Cell of origin | |  |  | |
| Non-GCB | | 21 | 44.7% | |
| GCB | | 25 | 53.2% | |
| Undetermined | | 1 | 2.1% | |
| IPI | |  |  | |
| 0-1 | | 26 | 55.3% | |
| 2-3 | | 14 | 29.8% | |
| 4-5 | | 6 | 12.8% | |
| Undetermined | | 1 | 2.1% | |
| Double-hit lymphoma | |  |  | |
| Yes | | 2 | 4.3% | |
| No | | 45 | 95.7% | |
| Relase status | |  |  | |
| Non-relapse | | 29 | 61.7% | |
| Relapse | | 16 | 34.0% | |
| Unclassified | | 2 | 4.3% | |
| international prognostic index, IPI; geminal center B-cell like, GCB. | | | | |
| Supplementary Table S2. Gene signatures characteristic of Macrophages, T cells, and B cells | | | | |
| Macrophages | T cells | | | B cells |
| CD68 | CD3D | | | MS4A1 |
| CD163 | CD3E | | | CD79A |
| CD14 | UBASH3A | | | CD79B |
| CSF1R | CD2 | | | CD19 |
|  | TRBC2 | | | PAX5 |

| Supplementary Table S3. The gene lists of macrophage signatures | | | | | |
| --- | --- | --- | --- | --- | --- |
| LZ VS DZ | | RLT VS DLBCL | | Non-relapse VS Relapse | |
| MacroSig3 (LZ) | MacroSig4 (DZ) | MacroSig5 (RLT) | MacroSig6 (DLBCL) | MacroSig7 (No relapse) | MacroSig8 (Relapse) |
| FOXE3 | LTBP1 | CR2 | C1QB | SELENOP | CCL22 |
| BCL2A1 | TTC9C | FCAMR | C1QC | C7orf25 | CA7 |
| HSPA1A | SLC16A7 | FDCSP | FGL2 | PITX1 | P3H4 |
| HAUS5 | GAB3 | CXCL14 | C1QA | TP53INP1 | DAAM1 |
| CLU | RRAGC | IGHG1 | PILRA | ZBTB38 | SNAPC1 |
| LMO2 | TMEM60 | IGHG3 | CCL4L2 | SDC3 | VEGFA |
| CHI3L1 | ZNF440 | IGHG4 | TAP1 | DONSON | STEAP1 |
| FCAMR | CCR7 | IGHG2 | CXCL9 | OSBPL3 | DRGX |
| PTPN2 | TCL1A | CLU | TYMP | ABCA2 | TRAM2 |
| IFNA14 | UBN2 | DSP | CSGALNACT2 | EPB41L2 | FITM1 |
| IDE | CDK13 | SERPINE2 | DUSP3 | PGM2L1 | RTKN2 |
| JUNB | LMAN2L | CDC42EP4 | EMILIN2 | IGHG1 | SLX4IP |
| PGAP6 | CIBAR2 | H2BC6 | SGTB | ATP8A1 | TEAD2 |
| CD83 | BDP1 | H4C6 | GBP1 | DYRK2 | H1-1 |
| FDCSP | C6 | FUCA1 | CSF3R | IGF1 | HSF5 |
| ZCWPW1 | KLRC4 | IGFBP2 | CLMN | TSPYL4 | RDX |
| SOGA1 | VPS37D | ELL3 | MCTP1 | TBC1D4 | SDK2 |
| ZSCAN32 | BTBD6 | MYBL1 | SERPINA1 | P2RX1 | RIBC1 |
| BTBD10 | PRC1 | UBE2J1 | FUOM | DNLZ | NEB |
| RAMP3 | EIF2AK2 | HMCES | SOD2 | A2M | REM2 |
| TXNRD1 | TSPAN15 | EPHX2 | LILRB2 | C3 | ECE1 |
| RANGRF | RNASE1 | NLK | SERPING1 | ADAP1 | RILPL2 |
| SRGN | GNE | ITIH5 | FCGR2C | LIPA | JAG1 |
| SCARA3 | METTL5 | ASB13 | STAT1 | CFD | KIAA1614 |
| RAB36 | FXR2 | EPAS1 | KDSR | ITGB7 | CXADR |
| TMEM184C | CHRNE | ATP6V0D2 | ENTPD6 | FOLR2 | HOMER2 |
| ZNF518B | ZFYVE21 | LPP | VAMP5 | TAF9B | EFCAB3 |
| ACHE | GPR174 | POU2AF1 | IL10RA | ZNF711 | TAS1R3 |
| SDAD1 | RUBCNL | P2RY8 | CEBPB | QPRT | GOLGA6A |
| RPUSD3 | TNFSF8 | NUGGC | WARS1 | KCNJ5 | S1PR3 |
| TMEM39B | SH3PXD2B | MATN2 | PDE4B | PIF1 | STAM |
| HNRNPLL | SYNE3 | SNX22 | PDLIM5 | SLC40A1 | KCNG4 |
| NECAP1 | PPP1R35 | MAML3 | ZEB2 | ELMO3 | RNF186 |
| TTI1 | SAP30L | CHN1 | NMRAL1 | SIGLEC8 | MT3 |
| NOP16 | C1QB | DEF8 | PRR14 | NUF2 | ASDURF |
| FAM207A | H1-1 | SCARA3 | CCR5 | ASB2 | STRADB |
| CR2 | TMEM250 | STK17B | IL17RA | MARCKSL1 | PKDCC |
| NSD1 | MELK | JCHAIN | UBE2L6 | SNED1 | KRTAP10-1 |
| BIRC3 | NUDT9 | SERPINF1 | PPRC1 | SFRP2 | HSD3B2 |
| NAA60 | PRR11 | MARCKSL1 | PFKP | TOX | HPDL |
| CHIT1 | C1QA | CD22 | SCO2 | RNASE1 | CHST1 |
| FBXO33 | C1QC | EZR | GBP5 | GPR34 | OTX1 |
| BICDL1 | MPZL2 | ITGB4 | TRIM65 | UCHL1 | CCR7 |
| SHPK | RAD51 | DERL3 | ITPR2 | CST1 | KRTAP5-10 |
| SULT2B1 | TNS4 | HMGN2 | HSPA6 | ITGAD | CPLANE1 |
| TMEM35B | TRIM68 | BCL7A | RIPOR1 |  | TRIM64B |
| CNN3 | CASK | RGS9 | FOS |  | ZNF570 |
| TAPT1 | ITGAD | S1PR2 | PAK1 |  | TGIF1 |
| MT1X | YJEFN3 | CRIP2 | RILPL2 |  | MMUT |
| SOX8 | H2BU1 | TOB2 | FTH1 |  | UBAP1L |
| NAA16 | OSBPL10 | RAB42 | GASK1B |  | TMTC4 |
| SOD2 | ZGRF1 | EPS15 | APOL3 |  | TTN |
| DNTTIP2 | TRIM62 | CUL3 | APOL1 |  | MT1H |
| PRKCH | PLP2 | H3C1 | CCL2 |  | KCNN4 |
| EFR3A | ADGRB1 | TSPAN10 | SLC1A3 |  | RHCE |
| ATP5MGL | DVL2 | TIMD4 | ARMC6 |  | LY6H |
| DBT | CSF1 | TERF2 | ALDH1A1 |  | ELFN1 |
| ITGB3BP | SYNJ2BP | RASSF2 | C1orf162 |  | TPRG1 |
| NPPC | TIMM21 | IGF1 | FLVCR1 |  | FAT3 |
| SLC19A2 | COL4A2 | SEL1L3 | SOAT1 |  | RASGRP2 |
| KIF7 | SLC44A2 | BPNT1 | ST3GAL5 |  | RNF224 |
| LRMDA | DMC1 | SLC30A4 | PSME2 |  | GK |
| UBA5 | CNDP2 | SPRED2 | PPP1R15A | | TGM5 |
| C20orf96 | APOF | DNAJC10 | FAM20C |  | NLRP6 |
| ATP13A5 | PLXNB1 | IRAG2 | PLEKHO1 |  | CLEC17A |
| CXCL13 | RAET1G | PAG1 | JAML |  | GGTLC2 |
| ILDR1 | SHISA7 | TCF7 | IRF1 |  | CPE |
| MARCHF8 | RBMX2 | RARRES1 | URB1 |  | WFDC2 |
| GGT5 | GSTZ1 | SIAH2 | OSCAR |  | JADE3 |
| CLCF1 | FBXO22 | RARRES2 | CCL5 |  | UBTFL1 |
| DHRS9 | TTPAL | DCK | ATG101 |  | MICU2 |
| IPO11 | GALC | SIGLEC8 | FAS |  | KAZALD1 |
| BTRC | MFSD12 | RAB6B | USP32 |  | AUNIP |
| PIP4P1 | KPNA2 | COLQ | CD163 |  | CCDC43 |
| MPG | CYP2S1 | CPNE5 | PRRT3 |  | ADAMTS14 |
| C10orf90 | PGM3 | HMGN1 | LILRB3 |  | ZNF444 |
| ICOS | CMTM5 | S1PR3 | IFITM3 |  | ZNF154 |
| SOX9 | RAB26 | MYO1E | S100A9 |  | ACSM3 |
| KIAA2026 | TMEM101 | AMFR | GRAMD1A | | CDX2 |
| ZNF35 | STAB1 | GNG4 | FGR |  | AOC2 |
| CDC42EP4 | CR1L | SOX8 | MANBA |  | ZNF268 |
| KRCC1 | RNF6 | LAMC2 | LRRK2 |  | RADX |
| STX11 | CD163L1 | NPHS1 | TAZ |  | SFTPB |
| VN1R4 | SLC25A43 | H2AC16 | KPTN |  | C10orf95 |
| ZNF484 | FAM151A | CSTA | GIMAP6 |  | RNF24 |
| CMTM2 | PDE6G | SUSD3 | PGD |  | OR5K1 |
| PRLHR | TTK | H2BC9 | MT2A |  | PIM1 |
| EPAS1 | ARHGEF39 | PEX5 | ABCC1 |  | LIMD1 |
| PYGB | AXL | H4C13 | IRAK3 |  | STK32B |
| SMCO4 | ZNF317 | TK1 | EPDR1 |  | PRSS58 |
| MEGF9 | KPNA1 | COL6A1 | JUNB |  | CLU |
| DNAJB11 | WDR89 | ROBO1 | IQSEC1 |  | CATSPERZ |
| ZFP3 | PILRA | CRIP1 | PTPDC1 |  | ATP4A |
| TNFRSF1B | DYNC2I2 | LY86 | ARMC2 |  | OR5D16 |
| LAP3 | WDR91 | ABHD12 | CBR1 |  | OR2D2 |
| STX6 | ZADH2 | SHCBP1 | HDHD5 |  | GUCY2F |
| C3 | AP3M2 | PLEKHF2 | ELOF1 |  | SYT5 |
| MAP7D1 | MIER2 | RANBP17 | HOXC4 |  | P2RY1 |
| ZNF516 | SLC39A13 | FAM174B | IQGAP2 |  | AJAP1 |
| THAP4 | CTXN1 | POLD4 | USHBP1 |  | ADAM33 |
| HTR3A | TYW1 | VPREB3 | APOBEC3G | | SLC22A31 |
| TRIM35 | TOP3B | IGSF10 | MGAT1 |  | CHRM1 |
| FSTL4 | BYSL | SYVN1 | ARMC10 |  | GCM2 |
| TSPAN6 | HELZ | SPARC | TP63 |  | ARMC3 |
| KLHL11 | HMGB2 | NAA38 | CRIM1 |  | HS6ST3 |
| SYT2 | MGST2 | TNFRSF21 | PTPRJ |  | CHRNB4 |
| PFAS | FRMD4B | DDR2 | HAPLN3 |  | SLC6A8 |
| ALG3 | ZFYVE19 | FAM76B | TRMT1 |  | ZNF616 |
| MYBBP1A | PPID | H1-3 | IFI27 |  | HCN3 |
| PLEK | MS4A6A | TSPAN13 | VAV2 |  | SOX11 |
| NIN | PLPP5 | QPRT | EEF2K |  | USP30 |
| LILRB3 | KLRB1 | CACNG5 | C4BPB |  | MT1A |
| FANCF | NENF | H4C1 | NOD1 |  | PRY |
| ZNF701 | PATJ | GGA2 | CD96 |  | FGF14 |
| TRPM2 | LHFPL6 | COL6A2 | SH3BP1 |  | MMRN2 |
| BARX2 | SNAP47 | PARP1 | POLR3D |  | SH3PXD2B |
| PCYOX1L | PNPLA4 | PNOC | EMILIN1 |  | UST |
| YARS1 | PLEKHA5 | MALT1 | KLHL8 |  | OR9Q1 |
| XPO5 | BACH2 | H3C7 | IER3 |  | SLC41A1 |
| CMPK1 | SLC35D1 | FBXO44 | ARHGEF10L | | SDC2 |
| ZNF593 | ITGB5 | BBS5 | RGS2 |  | C6orf163 |
| ARNTL2 | ALG10 | CMPK1 | FCGR3A |  | GALNS |
| ATAD2B | DHRS3 | SYNM | SSBP4 |  | ZNF689 |
| NPAS4 | ADORA3 | NUAK1 | ICAM1 |  | ZP1 |
| GMPPB | H3-2 | PTPRS | CSNK2A3 |  | ABHD17C |
| CDH1 | CDC20 | SMAD9 | DCLK2 |  | CLECL1 |
| IRF6 | ODF2L | MOB3A | LILRA5 |  | OR5L1 |
| POLR1F | PLEKHB1 | HMGB1 | ACACA |  | B3GAT1 |
| CHST11 | MAMDC2 | ICA1 | CD14 |  | SLC2A14 |
| NFKBIE | PACRG | ITPKB | TIMP1 |  | USP17L7 |
| PIK3R4 | HLX | VNN2 | SIK1 |  | RAB15 |
| LAX1 | ZNF100 | HDAC9 | ADGRE5 |  | BANK1 |
| SLC26A11 | SOX5 | H2BC3 | CCL3 |  | ATP6V1E2 |
| SLC7A5 | PHACTR4 | STAT5B | VPS72 |  | TUBB6 |
| PEX26 | GRK5 | PRLHR | GOLGA8J |  | CYB5R2 |
| KCNH4 | DYNLT3 | CCNG2 | GYG1 |  | P2RY14 |
| HSD3B7 | TRNAU1AP | IL21 | TMEFF1 |  | STEAP3 |
| TBC1D2 | SSPOP | USP34 | ZNF638 |  | CCND2 |
| MRTFB | SNTA1 | WIPF1 | SLC2A3 |  | SGSM1 |
| VPS54 | ETAA1 | USP7 | SLC31A2 |  | PTPN1 |
| LCE2D | ERCC1 | SERPINA9 | C6orf223 |  | LSAMP |
| DSCAM | PMM1 | FCHO1 | TRMU |  | PRSS12 |
| SKAP1 | C10orf95 | MFGE8 | SNX27 |  | TTC28 |
| BMT2 | PRR29 | FOXF1 | IFIT5 |  | ZFPM1 |
| EGR3 | RIN3 | LY75 | ALDH3B1 |  | GUCA1ANB |
| WDR41 | ABCB4 | RFTN1 | RHOBTB3 | | NCR2 |
| PARM1 | KIAA1522 | IGHA1 | ACBD6 |  | JHY |
| RPL22L1 | KLHDC1 | FGD6 | NINJ1 |  | CYP27B1 |
| CBR1 | VSIG8 | SCARB1 | THEMIS2 |  | POU4F1 |
| FN1 | PDCD11 | CARD14 | AKIP1 |  | MT1E |
| ERICH1 | MS4A14 | NSMF | YBX3 |  | CLEC4E |
| UBFD1 | POU6F1 | ZBTB38 | SHTN1 |  | CEMIP |
| LMTK3 | ZFAND2B | ERMARD | CHST7 |  | TERT |
| SLC9A1 | MLST8 | ANTXR1 | BST2 |  | IL36RN |
| RGMB | SCYL3 | VCAM1 | PSMG1 |  | IGSF9B |
| LAT | ALG14 | SGPL1 | ZDHHC23 |  | EFCAB12 |
| TNFRSF9 | NKX1-1 | TMEM131L | CFLAR |  | CRYZ |
| TRIP6 | KMT2A | CAMK1 | SLC39A14 |  | G0S2 |
| BAHD1 | MND1 | CCNB2 | ZC3HAV1L | | DGKE |
| N4BP2 | DAG1 | RDH10 | DOK2 |  | C1QL4 |
| ACER3 | S100A10 | MAP4K4 | TLR8 |  | AQP9 |
| CCDC102A | AK1 | SLC38A9 | MZT2B |  | KRTAP8-1 |
| PSPC1 | TMEM154 | IGKC | GZMB |  | S100A8 |
| PFDN4 | SMUG1 | SELENOP | GK |  | HNF4A |
| LTA | KEL | FAM107B | TTL |  | IL1R2 |
| TMEM171 | CCDC122 | CLPTM1 | DDX59 |  | MT1G |
| ZNF181 | TOR1A | NSRP1 | PIP5K1C |  | PDLIM1 |
| ARSA | ENDOG | HS3ST1 | HSPB1 |  | PRSS21 |
| BICRA | SLC27A4 | TIFA | TYROBP |  | ST18 |
| TRPM7 | SAMD4A | TMEM119 | NSDHL |  | TOP1MT |
| SLC6A8 | TBC1D10C | CD163L1 | CCBE1 |  | TRH |
| EDC4 | EVA1B | MEF2C | TGFB1 |  | CHST2 |
| EGR1 | TRIM33 | H3C3 | ACE |  | DEFB103A |
| GCA | CTSF | RBBP4 | GBP4 |  | SPACA6 |
| UBXN2B | TJAP1 | H1-1 | DIAPH2 |  | ZC3H12A |
| TRMT6 | PLXDC2 | HOPX | SNTB1 |  | EREG |
| HLA-G | TDRD7 | SYNE2 |  |  | CD44 |
| MAF | PKN2 | ZFAND4 |  |  | CRTAM |
| CRIP2 | CARMIL2 | DNASE1 |  |  | ROPN1B |
| UVSSA | CUL7 | SSBP2 |  |  | IGHM |
| GRAMD4 | PRDM10 | VAMP1 |  |  | H2AB1 |
| OTUD7A | ZNF394 | SSBP3 |  |  | ZBTB32 |
| ADCY3 | DIS3L2 | ADGRG5 |  |  | HIPK4 |
| DUSP2 | MPO |  |  |  | CXCL8 |
| ASMTL | CALHM2 |  |  |  | BLNK |
| TTLL5 | RILP |  |  |  | MST1 |
| TESPA1 | KIAA1109 |  |  |  | ZNF793 |
| NOL11 | DTX4 |  |  |  | GAB1 |
| MIER1 | STYX |  |  |  | IL7R |
| CBLC | PLCXD1 |  |  |  | MT1HL1 |
| C1S | XXYLT1 |  |  |  | UCMA |
| RRS1 | TSEN2 |  |  |  | MYO18B |
| CPD | NIBAN2 |  |  |  | MTURN |
| ATG2A | OR10H3 |  |  |  | ALDH6A1 |
| GORASP1 | SOX3 |  |  |  | PNMA5 |
| MICOS13 | TAF8 |  |  |  | BCL2 |
| IL32 | DUOXA1 |  |  |  | TLE1 |
| NDUFAF4 | TFAP2C |  |  |  | PKP3 |
| FBXW8 | TGFBI |  |  |  | FCGR2B |
| FADS2 | PLEKHD1 |  |  |  | IRF4 |
| SFSWAP | NKAPD1 |  |  |  | TNFRSF13B |
| LGALS8 | VAMP4 |  |  |  | DDIT4 |
| ADAT3 | SYTL3 |  |  |  | PLAUR |
| DNPH1 | RUNX1T1 |  |  |  | H2AC4 |
| LZIC | SCO1 |  |  |  | CD300E |
| ZXDB | HUS1 |  |  |  | CXCL3 |
| MAGT1 | CCNB2 |  |  |  | SLC2A3 |
| SPG11 | CENPE |  |  |  | HSD11B1 |
| DPY19L1 | SLF1 |  |  |  | CXCL5 |
| LANCL2 | TNK1 |  |  |  | SLC39A8 |
| SLC35F6 | CENPO |  |  |  | PDGFB |
| PPP1R13B | DGKQ |  |  |  | L1CAM |
| AFAP1 | GHDC |  |  |  |  |
| TENT4B | THOC7 |  |  |  |  |
| USF3 | FAM111B |  |  |  |  |
| METTL3 | IKBKB |  |  |  |  |
| EPHX2 | PODXL |  |  |  |  |
| AFAP1L2 | DNAH8 |  |  |  |  |
| SIK3 | ESRRA |  |  |  |  |
| MRPS33 | CA3 |  |  |  |  |
| RO60 | DBR1 |  |  |  |  |
| ZNF136 | FOXO6 |  |  |  |  |
| RDH14 | ZFP57 |  |  |  |  |
| KCTD10 | EIF4EBP1 |  |  |  |  |
| PTGR1 | KCTD3 |  |  |  |  |
| CDC42EP3 | POP7 |  |  |  |  |
| CD226 | NFKBIB |  |  |  |  |
| VARS2 | RETREG2 |  |  |  |  |
| TRIP10 | BSPRY |  |  |  |  |
| EXOC3L4 | ODF3B |  |  |  |  |
| FUCA2 | COL6A3 |  |  |  |  |
| MALL | SPRR2E |  |  |  |  |
| SYNM | DLG3 |  |  |  |  |
| ZNF235 | SLC22A2 |  |  |  |  |
| EFHD2 | GTPBP3 |  |  |  |  |
| RIC8A | RBMS2 |  |  |  |  |
| IMMT | TSEN15 |  |  |  |  |
| SLC35A5 | DENND11 |  |  |  |  |
| CEP135 | STRN |  |  |  |  |
| TIGIT | DEXI |  |  |  |  |
| PTPRF | SCX |  |  |  |  |
| GRPEL1 | SAV1 |  |  |  |  |
| CXCL16 | TMEM116 |  |  |  |  |
| SKAP2 | PLA2G15 |  |  |  |  |
| FBXO6 | STK26 |  |  |  |  |
| GSDMB | APC |  |  |  |  |
| EPHX1 | FES |  |  |  |  |
| NDST1 | C21orf58 |  |  |  |  |
| CCNQ | MTG2 |  |  |  |  |
| ZBED6CL | SCUBE2 |  |  |  |  |
| XRN2 | TM2D1 |  |  |  |  |
| JMJD4 |  |  |  |  |  |
| PARVB |  |  |  |  |  |
| LEMD2 |  |  |  |  |  |
| AP1G2 |  |  |  |  |  |
| IRF4 |  |  |  |  |  |
| PLA2G3 |  |  |  |  |  |
| FCER2 |  |  |  |  |  |
| ACO1 |  |  |  |  |  |
| FKRP |  |  |  |  |  |
| PIGS |  |  |  |  |  |
| CTNND1 |  |  |  |  |  |
| TRIM13 |  |  |  |  |  |
| EGFLAM |  |  |  |  |  |
| UBA7 |  |  |  |  |  |
| MALT1 |  |  |  |  |  |

| Supplementary Table S4. HR and 95% CI of overall survival in DLBCL patients after 6 cycles of R-CHOP | | | |
| --- | --- | --- | --- |
| Groups | Datasets | HR | 95% CI |
| MacroSig6 (DLBCL) versus MacroSig5 (RLT) | Len et al. (*N* = 420) | 2.06 | 1.38-3.07 |
|  | Dubois et al. (*N* = 223) | 2.51 | 1.14-5.51 |
|  | Reddy et al. (*N* = 773) | 1.43 | 1.03-1.99 |
|  | Schmitz et al. (*N* = 481) | 1.95 | 1.18-3.21 |
|  | Lacy et al. (*N* = 1,149) | 1.37 | 1.13-1.67 |
| MacroSig8 (Relapse) versus MacroSig7 (Non-Relapse) | Len et al. (*N* = 420) | 1.97 | 1.34-2.9 |
|  | Visco et al. (*N* = 498) | 1.54 | 1.06-2.23 |
|  | Reddy et al. (*N* = 773) | 1.52 | 1.1-2.09 |
|  | Schmitz et al. (*N* = 481) | 2.52 | 1.57-4.04 |
|  | Lacy et al. (*N* = 1,149) | 1.5 | 1.23-1.83 |
| MacroSig4 (DZ) versus MacroSig4 (LZ) | Len et al. (*N* = 420) | 2.38 | 1.61-3.53 |
|  | Visco et al. (*N* = 498) | 1.47 | 1.01-2.14 |
|  | Dubois et al. (*N* = 223) | 2.15 | 1.07-4.32 |
|  | Reddy et al. (*N* = 773) | 1.53 | 1.13-2.08 |
|  | Chapuy et al. (*N* = 137) | 2.87 | 1.12-7.33 |
|  | Schmitz et al. (*N* = 481) | 2.43 | 1.5-3.94 |
|  | Sha et al. (*N* = 913) | 2.03 | 1.36-3.05 |
|  | Lacy et al. (*N* = 1,149) | 1.55 | 1.29-1.88 |
| diffuse large B-cell lymphoma, DLBCL; rituximab with cyclophosphamide, doxorubicin, vincristine, and  prednisolone (R-CHOP); macrophage signatures, MacroSigs; reactive lymphoid tissue, RLT; confidence interval, CI; light zone, LZ; dark zone, DZ. | | | |

| Supplementary Table S5. Multivariable analysis of MacroSig4 (DZ) adjust for IPI scores* | | | | |
| --- | --- | --- | --- | --- |
| Dataset | Variable | OS | | |
|  |  | HR | 95% CI | *P* value |
| Dubois (*N* = 223) | MacroSig4 (DZ) | 2.03 | 1.01-4.08 | 0.048 |
| Reddy (*N* = 773) | MacroSig4 (DZ) | 1.23 | 0.87-1.73 | 0.25 |
| Chapuy (*N* = 137) | MacroSig4 (DZ) | 2.68 | 1.04-6.89 | 0.041 |
| Schmitz (*N* =481) | MacroSig4 (DZ) | 2.06 | 1.19-3.55 | 0.0095 |
| Sha (*N* = 913) | MacroSig4 (DZ) | 1.93 | 1.28-2.91 | 0.0017 |
| Lacy (*N* = 1,149) | MacroSig4 (DZ) | 1.31 | 1.03-1.68 | 0.029 |
| *The IPI scores of Len et al. and Visco et al. were not available. Dark zone, DZ; international prognostic index, IPI; overall survival, OS; hazard ratio, HR;  confidence interval, CI. | | | | |

### Supplementary Figures

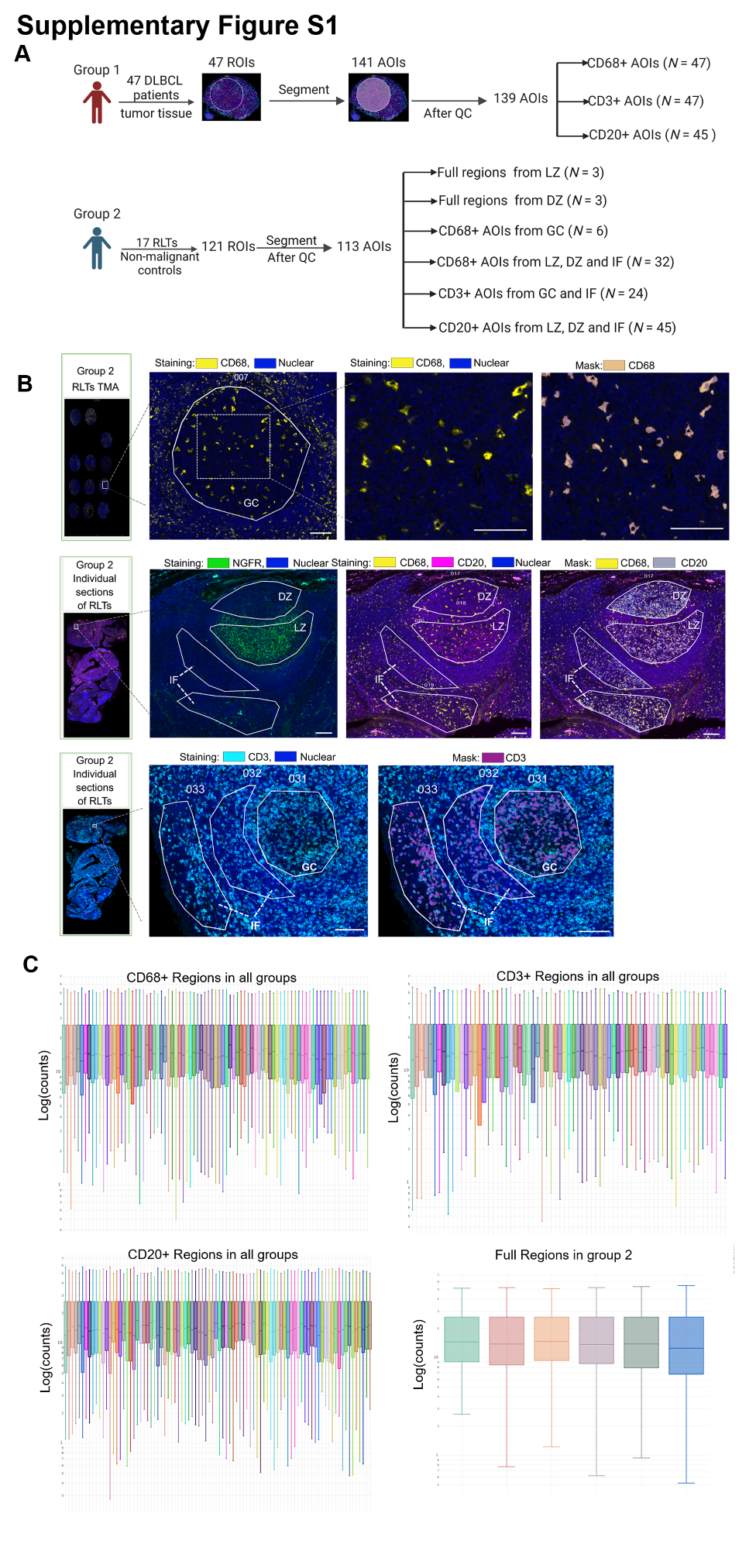

**Supplementary Figure S1. DSP workflow, mask segmentations and normalization of count data. (A)** Schematic of study groups. **(B)** Immunofluorescence staining of RLTs. In RLTs TMA of Group 2, CD68 stained macrophages (yellow), and SYTO 13 stains nuclei (blue). In individual tonsil sections of Group 2, NGFR illuminates light zone (green), CD68 stains macrophages (yellow), CD3 stains T cells (cyan), CD20 stains B cells (magenta), and SYTO 13 stains nuclei (blue). After ROIs selection, each cell type was segmented by their corresponding masks. Representative images were shown. Scale bar: 100 μm. **(C)** Violin plot displayed the RNA counts of each sample in all regions after Q3 normalization. digital spatial profiling, DSP; reactive lymphoid tissues, RLTs; tissue microarray, TMA; regions of interest, ROI; areas of interest, AOI; nerve growth factor receptor, NGFR; light zone, LZ; dark zone, DZ; inter-follicular, IF, germinal center, GC.

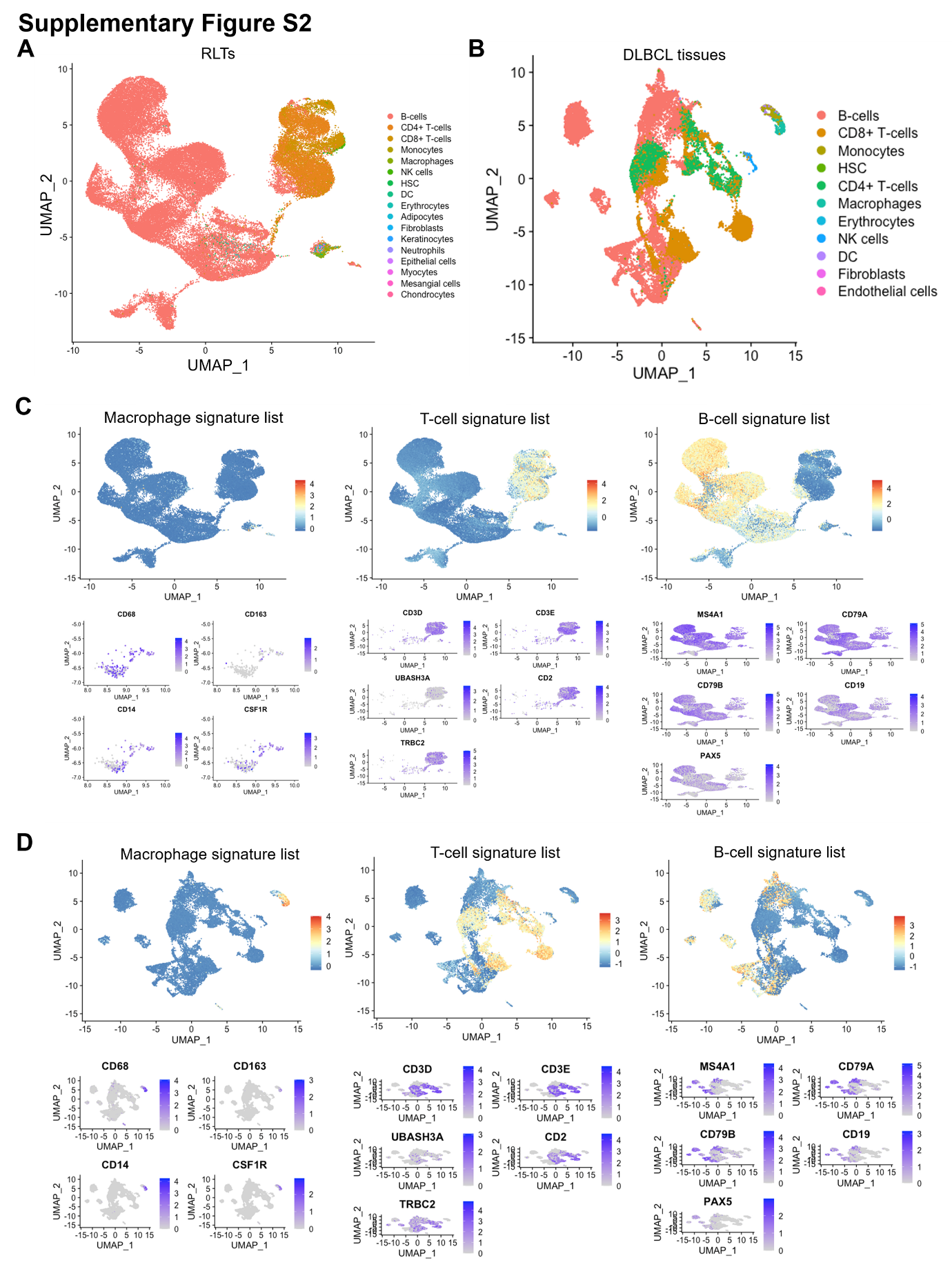

**Supplementary Figure S2. The signatures of macrophage, T cell, and B cell were verified using scRNA-seq data. (A)** UMAP of RLTs profiled in public scRNA-seq datasets (*N* = 10). **(B)** UMAP of DLBCL tissues profiled in public scRNA-seq data (*N* = 10). **(C)** The gene signatures of macrophage, T cell, and B cell were projected to public scRNA-seq data of RLTs, respectively. **(D)** The signatures of macrophage, T cell, and B cell were projected to public scRNA-seq data of DLBCL tissues, respectively. diffuse large B-cell lymphoma; DLBCL; single cell sequencing, scRNA-seq; uniform manifold approximation and projection, UMAP; reactive lymphoid tissues, RLTs.

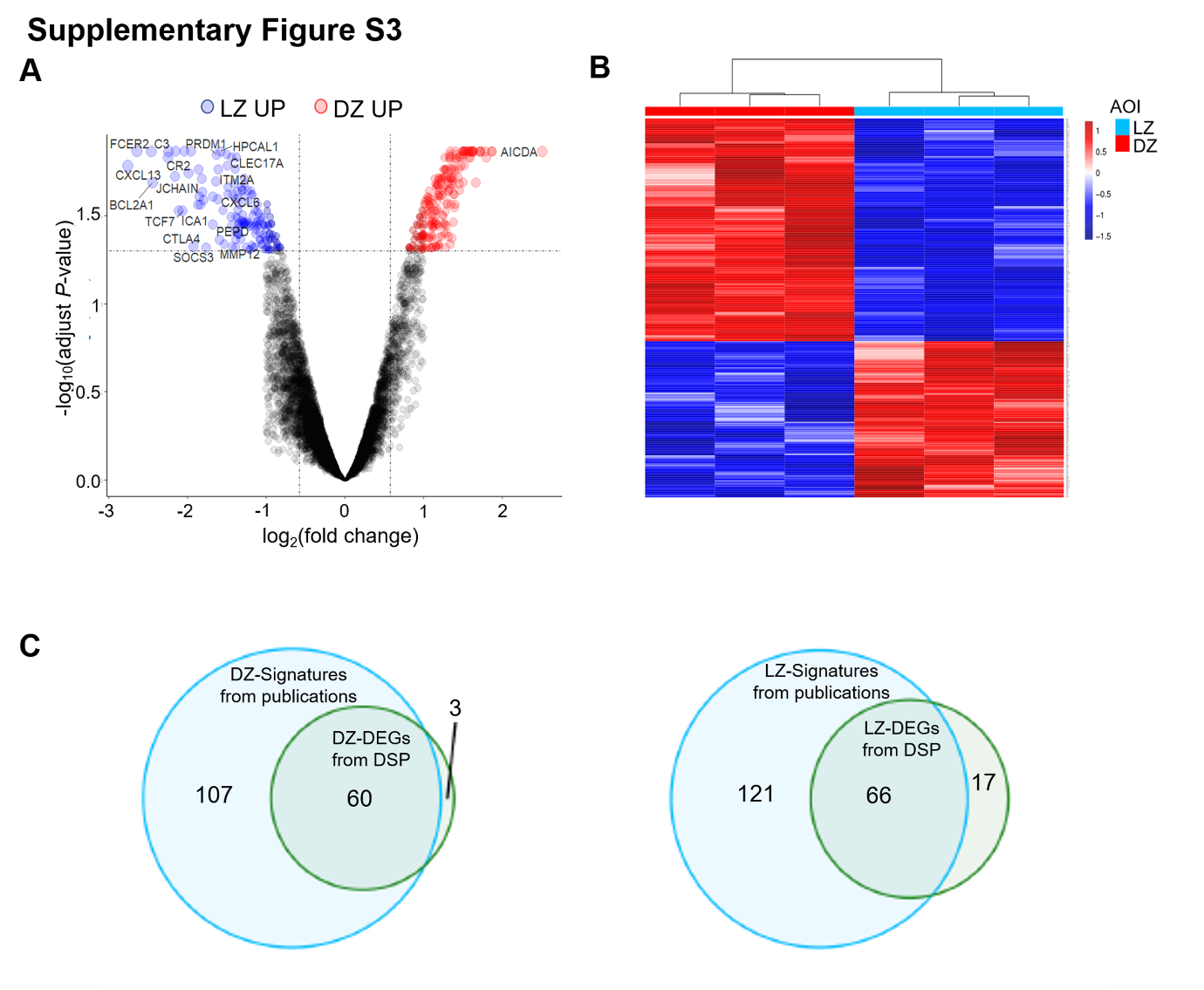

**Supplementary Figure S3. Validating of gene expression patterns in the LZ and DZ (A-B)** Volcano plot and heatmap showed the DEGs of full regions between LZ and DZ (*N* = 6). **(C)** Venn diagram displayed the overlapping DEGs from LZ and DZ between our data and previous publications. differentially expressed genes, DEGs.

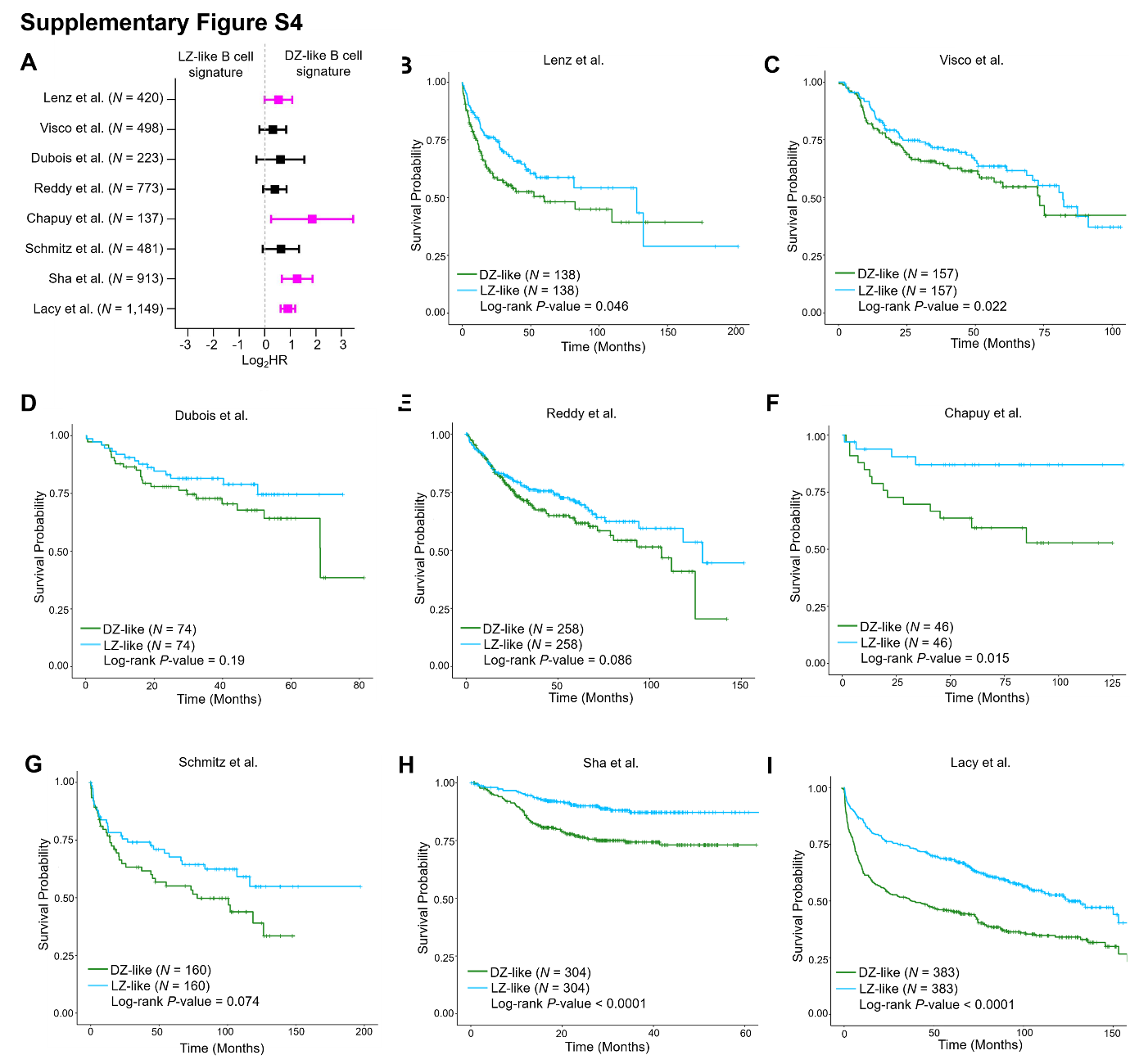

**Supplementary Figure S4. Evaluation of LZ- and DZ-like B-cell signatures as predictors of DLBCL patient prognosis. (A)** Univariate Cox proportional hazards model analysis for LZ- and DZ-like B cell signatures in eight publicly available DLBCL datasets. **(B-I)** Kaplan–Meier analyses showed that DZ-like B-cell signature was associated with poor OS in DLBCL patients using distinct DLBCL datasets.

| **Item to be reported** | | **Page no.** |
| --- | --- | --- |
| **INTRODUCTION** | |  |
| 1 | State the marker examined, the study objectives, and any pre-specified hypotheses. | Page 5 of main manuscript, |
| **MATERIALS AND METHODS** | |  |
| *Patients* | |  |
| 2 | Describe the characteristics (e.g., disease stage or co-morbidities) of the study patients, including their source and inclusion and exclusion criteria. | Page 5 of main manuscript, and Supplementary Table S1. |
| 3 | Describe treatments received and how chosen (e.g., randomized or rule-based). | Page 5 of main manuscript |
| *Specimen characteristics* | |  |
| 4 | Describe type of biological material used (including control samples) and methods of preservation and storage. | Page 5 of main manuscript |
| *Assay methods* | |  |
| 5 | Specify the assay method used and provide (or reference) a detailed protocol, including specific reagents or kits used, quality control procedures, reproducibility assessments, quantitation methods, and scoring and reporting protocols. Specify whether and how assays were performed blinded to the study endpoint. | Page 5-6 of main manuscript, and Supplementary methods |
| *Study design* | |  |
| 6 | State the method of case selection, including whether prospective or retrospective and whether stratification or matching (e.g., by stage of disease or age) was used. Specify the time period from which cases were taken, the end of the follow-up period, and the median follow-up time. | Page 5 of main manuscript, and Page 2 of supplementary data |
| 7 | Precisely define all clinical endpoints examined. | Page 2 of supplementary data |
| 8 | List all candidate variables initially examined or considered for inclusion in models. | Page 5 of main manuscript, Page 2 of supplementary data |
| 9 | Give rationale for sample size; if the study was designed to detect a specified effect size, give the target power and effect size. | Page 5 of main manuscript |
| *Statistical analysis methods* | |  |
| 10 | Specify all statistical methods, including details of any variable selection procedures and other model-building issues, how model assumptions were verified, and how missing data were handled. | Page 6 of main manuscript, and supplementary methods |
| 11 | Clarify how marker values were handled in the analyses; if relevant, describe methods used for cutpoint determination. | Page 4-5 of supplementary data |
| **RESULTS** | |  |
| *Data* | |  |
| 12 | Describe the flow of patients through the study, including the number of patients included in each stage of the analysis (a diagram may be helpful) and reasons for dropout. Specifically, both overall and for each subgroup extensively examined report the numbers of patients and the number of events. | Page 5 of main manuscript, and supplementary methods |
| 13 | Report distributions of basic demographic characteristics (at least age and sex), standard (disease-specific) prognostic variables, and tumor marker, including numbers of missing values. | Supplementary Table S1 and Supplementary Figure S1A |
| *Analysis and presentation* | |  |
| 14 | Show the relation of the marker to standard prognostic variables. | Figure 6-7, Supplementary Figure S4 |
| 15 | Present univariable analyses showing the relation between the marker and outcome, with the estimated effect (e.g., hazard ratio and survival probability). Preferably provide similar analyses for all other variables being analyzed. For the effect of a tumor marker on a time-to-event outcome, a Kaplan-Meier plot is recommended. | Figure 6-7, Supplementary Figure S4 |
| 16 | For key multivariable analyses, report estimated effects (e.g., hazard ratio) with confidence intervals for the marker and, at least for the final model, all other variables in the model. | Figure 6-7, Supplementary Figure S4 |
| 17 | Among reported results, provide estimated effects with confidence intervals from an analysis in which the marker and standard prognostic variables are included, regardless of their statistical significance. | Supplementary Table S4 |
| 18 | If done, report results of further investigations, such as checking assumptions, sensitivity analyses, and internal validation. | Page 7, 12 of main manuscript, Figure 1C-D, Supplementary Figure S2-S3 |
| **DISCUSSION** | |  |
| 19 | Interpret the results in the context of the pre-specified hypotheses and other relevant studies; include a discussion of limitations of the study. | Page 12-16 of main manuscript |
| 20 | Discuss implications for future research and clinical value. | Page 12-16 of main manuscript |
